# Th17.1 cell driven sarcoidosis-like inflammation after anti-BCMA CAR T cells in multiple myeloma

**DOI:** 10.1101/2022.12.08.22283148

**Authors:** Alexander M. Leipold, Rudolf A. Werner, Johannes Düll, Pius Jung, Mara John, Emilia Stanojkovska, Xiang Zhou, Hannah Hornburger, Anna Ruckdeschel, Oliver Dietrich, Fabian Imdahl, Tobias Krammer, Stefan Knop, Andreas Rosenwald, Andreas Buck, Leif Erik Sander, Hermann Einsele, K. Martin Kortüm, Antoine-Emmanuel Saliba, Leo Rasche

**Affiliations:** Helmholtz Institute for RNA-based Infection Research (HIRI), Helmholtz-Center for Infection Research (HZI), Würzburg, Germany; Mildred Scheel Early Career Center, University Hospital of Würzburg, Würzburg, Germany; Department of Nuclear Medicine, University Hospital Würzburg, Würzburg, Germany; Department of Internal Medicine 2, University Hospital of Würzburg, Würzburg, Germany; Department of Internal Medicine 1, University Hospital of Würzburg, Würzburg, Germany; Institute of Pathology, University of Würzburg, Würzburg, Germany; Charité - Universitätsmedizin Berlin, Department of Infectious Diseases and Respiratory Medicine, Charité, Universitätsmedizin Berlin, Berlin, Germany; German Center for Lung Research (DZL)

## Abstract

Pseudo-progression and flare-up phenomena constitute a novel diagnostic challenge in the follow-up of patients treated with immune-oncology drugs. We present a case study on pulmonary flare-up after Idecabtagen Vicleucel (Ide-cel), a BCMA targeting CAR T-cell therapy, and used single-cell RNA-seq (scRNA-seq) to identify a Th17.1 driven autoimmune mechanism as the biological underpinning of this phenomenon. By integrating datasets of various lung pathological conditions, we revealed transcriptomic similarities between post CAR T pulmonary lesions and sarcoidosis. Furthermore, we explored a noninvasive PET based diagnostic approach and showed that tracers binding to CXCR4 complement FDG PET imaging in this setting, allowing discrimination between immune-mediated changes and true relapse after CAR T-cell treatment. In conclusion, our study highlights a Th17.1 driven autoimmune phenomenon after CAR T, which may be misinterpreted as disease relapse, and that imaging with multiple PET tracers and scRNA-seq could help in this diagnostic dilemma.

## INTRODUCTION

CAR T-cell therapy is establishing itself as a new standard of care in relapsed and refractory multiple myeloma (RRMM), and the field is moving towards use of this modality in early treatment lines^1^. For example, anti-BCMA Ide-cel provides a median progression-free survival of 12.1 months in a heavily pretreated and triple-refractory patient population^2^. Yet, there are a number of open issues including the lack of data on optimal follow-up assessments and autoimmune phenomena post CAR T therapy. In this context it is worth mentioning that patients with autoimmune diseases were usually excluded from trials and the impact of these novel treatments on pre-existing conditions is unknown.

Appreciating the high incidence of extramedullary lesions and patchy disease patterns seen in the RRMM setting, whole body 18F-Fluorodeoxyglucose (FDG) PET/CT imaging is increasingly used in addition to serological M-protein testing and bone marrow aspirates for staging prior to and after CAR T infusion. Whether FDG is the best tracer in this setting has yet to be determined, as the technique is being based not only on the quantification of increased glucose uptake by tumor cells but also by others such as inflammatory cells^3^. Given the immunologic mode of action of CAR T-cell treatment, atypical response patterns related to CAR T-cell expansion and local inflammation could challenge image interpretation and subsequent response assessment. While data on MM have still to emerge, two pilot studies in lymphoma described pseudo-progression in patients after CD19 directed CAR T-cell therapy^4,5^. Pseudo-progression refers to an increase in the target lesion size due to influx of immune cells, and is a well-known event after checkpoint blockade therapy, seen in up to 10% of patients^6,7^. Other checkpoint inhibitor-related adverse events include sarcoidosis-like reactions, presenting as pulmonary nodules and mediastinal lymphadenopathy^8^. In general, the incidence of immune-related changes after CAR T therapy in MM is unknown, but strategies to discriminate progression from immune-related phenomena are required as these products become more commonly used.

Here, we report on a sarcoidosis-like flare-up after anti-BCMA CAR T-cell product Ide-cel and present our strategy to discriminate between autoimmune phenomena and true relapse using multiple PET tracers along with single-cell RNA-sequencing (scRNA-seq).

## RESULTS

### Case presentation

A patient in their 60’s was diagnosed with IgA kappa MM with osteolytic bone disease, anemia, and 80% bone marrow infiltration by malignant plasma cells. In addition, biopsy-proven extramedullary disease (EMD) was found located to both lungs, explaining months of violent coughing prior to presentation. Fluorescent in situ hybridization revealed high-risk cytogenetics including t(14;20), amp(1q), and del(1p). The patient responded to induction therapy using bortezomib, cyclophosphamide, dexamethasone, but stem cell harvest was skipped due to the COVID-19 pandemic. A bortezomib based bridging therapy failed, M-protein levels increased and salvage therapy was administered using daratumumab, pomalidomide, dexamethasone to which the patient was refractory. Given the limited options at this stage, the patient was treated with the anti-BCMA CAR T-cell product ide-cel. PET imaging using multiple tracers was conducted at baseline, 3-month follow-up, and 6-month follow-up (**Fig 1A-C, Supplementary Fig 1A-F**). Baseline FDG PET scan showed multiple focal bone lesions at the axial and appendicular skeleton as well as pulmonary manifestations with mediastinal lymphadenopathy (**Fig 1A, Supplementary Fig 1A**). The patient responded to carfilzomib-containing bridging therapy^9^, and the target dose of 450 × 10^6^ CAR T cells was infused. The patient experienced rapid-onset cytokine release syndrome (CRS) grade 1, which was successfully treated with repetitive doses of tocilizumab. Respiratory symptoms fully resolved during bridging therapy but came back 10 days after CAR T infusion. Disease assessment was performed at 3 months of follow-up revealing undetectable MRD in a bone marrow aspirate and full resolution of focal bone lesions but still disseminated FDG uptake at the lung and mediastinal lymph nodes. To narrow the differential diagnoses down, we performed a PET using ^68^Ga-DOTA(0)-Phe(1)-Tyr(3)-octreotide (^68^Ga-DOTATOC) that usually detects sarcoidosis with high sensitivity and specificity^10^, which was negative. Negative results were also seen for ^68^Ga-Pentixafor (binds to CXCR4), which is an alternative tracer to FDG in the detection of malignant plasma cells ^11^ (**Fig 1B, Supplementary Fig 1B-D**). Bronchoalveolar lavage (BAL) along with endobronchial ultrasound guided biopsy was done, showing a CD4 to CD8 ratio of 2.1 on cytology and noncaseating granuloma formation in the biopsy without evidence of residual MM cells. Infectious disease workup included microscopy, culture, ELISA and PCR for fungal and mycobacterial pathogens, which was negative.

**Figure 1:**
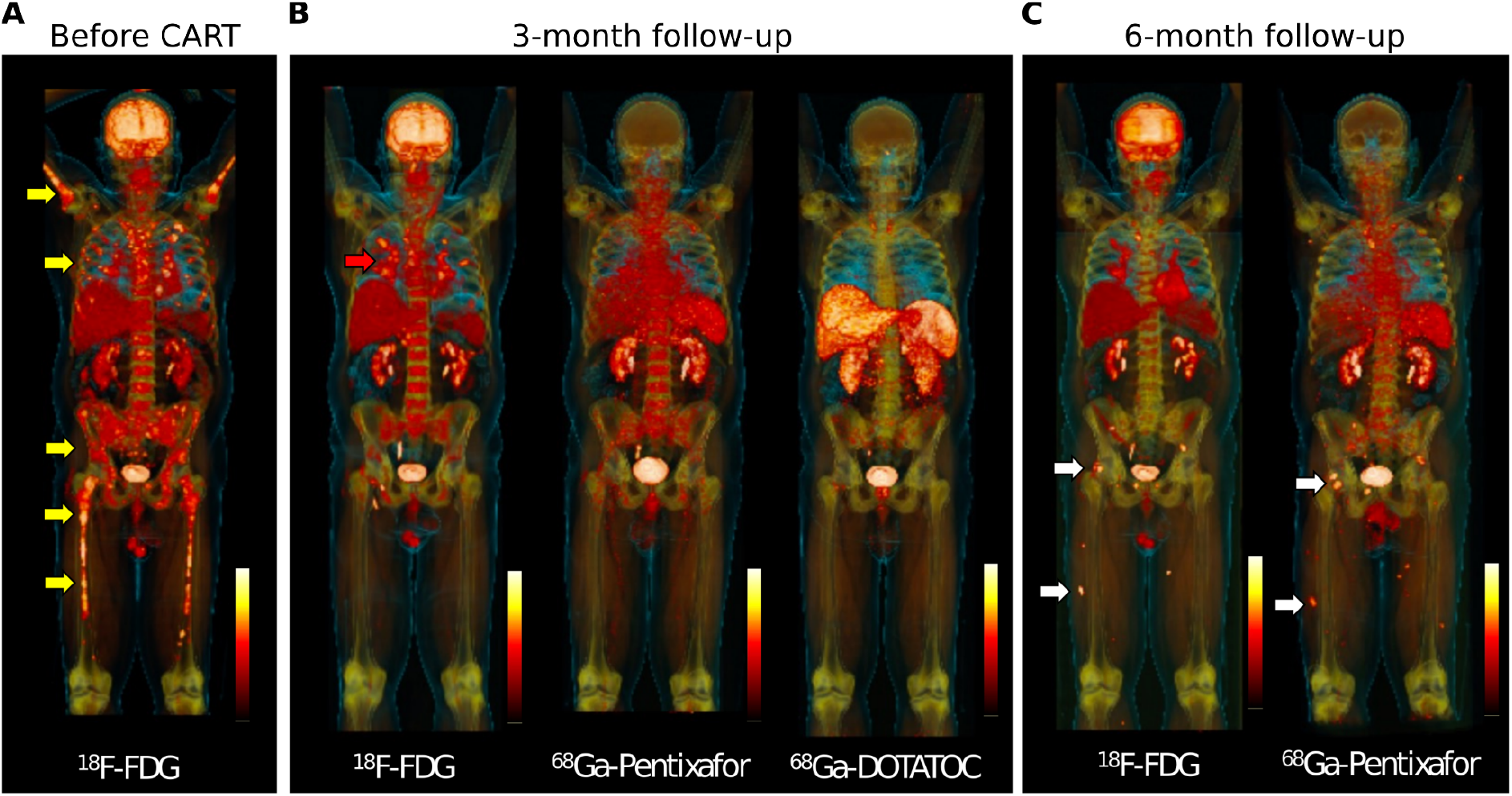
Whole body PET imaging at baseline and during follow-up using multiple PET tracers. **A**, FDG-PET at baseline shows multiple focal lesions at the axial and appendicular skeleton (yellow arrows). **B**, Follow-up assessment at 3-month after CAR T infusion shows full resolution of focal lesions, but residual FDG uptake located to the lung (red arrow). Matched-simultaneous CXCR4-targeted Pentixafor and DOTATOC PET did not show focal uptake. **C**, Matched-simultaneous FDG and CXCR4 PET 6 months after CAR T depicted new focal lesions at the pelvis and lower limbs in line with relapse (white arrows).

### Single-cell RNA-sequencing reveals presence of Th17.1 cells in the bronchoalveolar lavage

To unravel the biological underpinnings, we performed scRNA-seq on the BAL sample including 6,042 cells (henceforth called CART-BAL), of which none was identified as plasma cell (**Fig 2A, Supplementary Fig 2A-D, Supplementary Table 1**). Quality metrics were satisfactory across identified cell types (**Fig 2B**). As the CD4:CD8 T-cell ratio in both, cytology and scRNA-seq was increased (**Supplementary Fig 2E**), we decided to perform a focused transcriptomic analysis of T-cells. They were mainly CD4-positive cells exhibiting simultaneous expression of genes associated with Th1 T-cells (*TBX21, CXCR3, IFNG, TNF*) and genes associated with Th17 T-cells (*RORC, IL23R, CCR6*, CCL20, *KLRB1*), indicative for a Th1-polarized Th17 phenotype (**Fig 2C-E, Supplementary Fig 3A**,**B, Supplementary Table 2**). Furthermore, these cells expressed *IL4I1, ABCB1*, and *CSF2*^*12–14*^, while *IL17A, IL17B, IL17E* (*IL25*), and *IL17F* were not expressed (**Fig 2D**,**E, Supplementary Fig 3C**). Of note, the terminology for these Th1-polarized Th17 cells in the literature is not uniform. T-cell phenotypes with similar characteristics have been coined Th17.1, Th1/17, nonclassical Th1, and extinguish (ex)Th17^15–17^. They have been described to be pro-inflammatory and pathogenic and are implicated in several auto-immune diseases, including sarcoidosis^13,14,17–19^. To stay consistent with previous reports in pulmonary sarcoidosis, in which the majority of cells produced IFNγ but not IL17, we will refer to them as Th17.1 cells^17^. We also identified CD8-positive cells exhibiting gene expression resembling Th17.1 cells **(Fig 2C-E)**. Consequently, we termed this cluster CD8 Th17.1-like. Also CD8-positive cells with Th17-like features and plasticity towards *IFNG* expression have been linked to autoimmune diseases^20–23^. Th17.1 and CD8 Th17.1-like cells expressed a number of pro-inflammatory cytokines, including *IFNG, TNF, CSF2* (*GM-CSF*), *CCL20*, and *IL26*, highlighting their polyfunctional nature^15,24^ (**Fig 2D**,**E, Supplementary Fig 3C**). To validate the identified Th17.1 cells as genuine, we examined co-expression of four Th1-associated (*IFNG, TNF, TBX21, CXCR3*) and four Th17-associated (*CCL20, IL23R, RORC, CCR6*) genes in individual cells. They were considered co-expressing when at least one Th1- and one Th17-associated gene was detected in a single cell, and we confirmed co-expression in the majority of Th17.1 and CD8 Th17.1-like cells. (**Fig 2F**).

**Figure 2:**
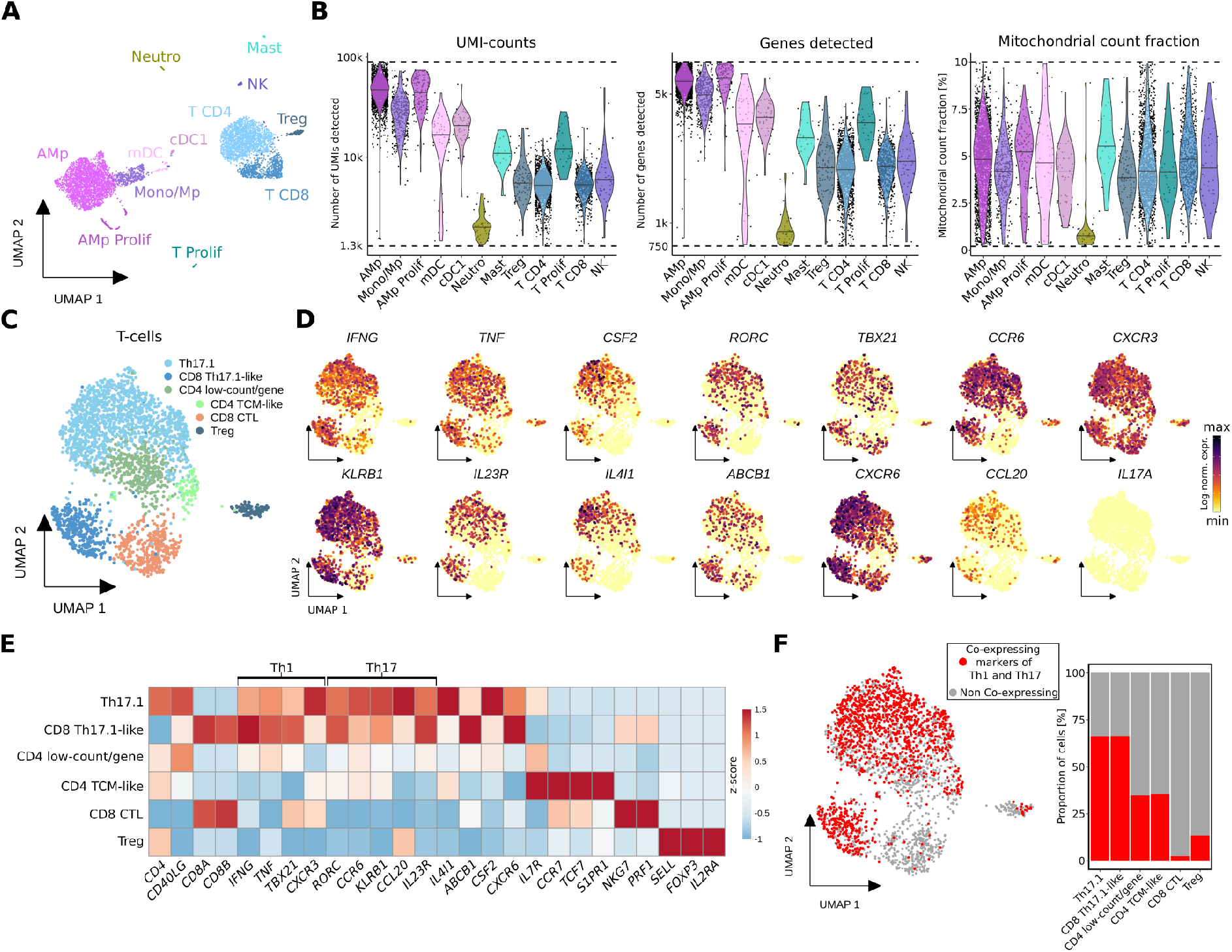
Single-cell analysis of bronchoalveolar lavage of a MM patient with FDG-avid pulmonary manifestation on PET-imaging suggests Th17.1-driven autoimmune phenomenon following anti-BCMA CAR T treatment. **A**, UMAP embedding of 6,042 single-cell transcriptomes from two technical replicates from BAL (bronchoalveolar lavage) of a MM patient at three months follow-up after CAR T therapy (CART-BAL). Cell type annotation was based on expression of canonical marker genes (**Supplementary Fig 2D**). **B**, Data quality metrics across cell types in CART-BAL depicted in violin plots. Dashed lines represent thresholds that were used for quality control filtering. For UMI-counts and genes detected, log10-scale is shown. Lines in violins show medians per cell type. **C**, UMAP embedding of 2,778 T-cells (T CD4, T CD8, Treg from **(C)**) from CART-BAL colored by subset annotation. **D**, Log-normalized gene expression of Th1-associated genes (*IFNG, TNF, TBX21, CXCR3*), Th17-associated genes (*RORC, CCR6, KLRB1, IL23R, CCL20*) and further selected genes (*CSF2, IL4I1, ABCB1, CXCR6*) that together characterize Th17.1-cells and *IL17A* color-coded and projected onto the UMAP embedding from (**C**). **E**, Heatmap showing the z-score of mean log-normalized expression of selected genes per T-cell subset identified in (**C**). **F**, Co-expression of Th1-associated (*IFNG, TNF, TBX21, CXCR3*) and Th17-associated (*CCL20, IL23R, RORC, CCR6*) genes in individual cells was assessed and projected onto the UMAP embedding as well as depicted as bar plot. Cells were deemed co-expressing when at least one Th1- and one Th17-associated gene was detected. Abbreviations: AMp - alveolar macrophage; Mono - monocyte; Mp - macrophage; CTL: cytotoxic T-lymphocyte; TCM - central memory T-cell.

### Transcriptional similarities between post CAR T pulmonary lesions and sarcoidosis

To further explore the role of Th17.1 cells in CART-BAL, we integrated the data with additional publicly available scRNA-seq datasets (**Fig 3A, Supplementary Fig 4A**,**B, Supplementary Table 1**). First, we used data from three healthy control BALs (HC in **Fig 3**), in order to confirm our findings to be truly related to pathological conditions^25^. Next, we used BAL data from seven patients suffering from severe COVID-19 acute respiratory distress syndrome (ARDS) with profound profibrotic lung remodeling (COVID-19 in **Fig 3**), constituting a control for overwhelming immune responses to an infectious trigger^26^. Last, we used BAL data from four pulmonary sarcoidosis patients (Sarc in **Fig 3**), as localisation of lesions and their appearance on FDG-PET was reminiscent of sarcoidosis^27^. We isolated and re-analysed T-cells and identified several subsets based on their gene expression profile, including Th17.1 cells, (**Fig 3B-D, Supplementary Fig 5A-C, Supplementary Fig 6A**,**B, Supplementary Table 2**). Th17.1 cells were validated using module scores built on two published gene signatures of Th1-polarized Th17 cells^28,29^ (**Fig 3E, Supplementary Fig 6C, Supplementary Table 1**). T-cells from sarcoidosis patients predominantly localized to the same UMAP space as CART-BAL T-cells, in line with profound phenotypic similarities (**Fig 3F**). To further explore the similarity between post CAR T pulmonary changes and sarcoidosis, we applied UniFrac distance calculation^30^. Here, a distance of 0 refers to a situation in which two conditions have exactly the same composition while a distance of 1 refers to completely different compositions. A UniFrac distance of 0.09 confirmed profound overlap between the two pathological conditions (**Fig 3G**). Of note, the module scores based on the two Th1-polarized Th17 gene signatures were high in sarcoidosis Th17.1 cells (**Supplementary Fig 6D**). To further interrogate similarities between CART-BAL and sarcoidosis T-cells, we projected skin derived single T-cell transcriptomes from three cutaneous sarcoidosis patients onto the integrated BAL T-cell embedding^31^ (**Supplementary Fig 7A-D**). Here, a substantial proportion of cells mapped onto the UMAP space of Th17.1 cells (**Supplementary Fig 7A**). As the mapping requires confirmation, we performed three more validation steps. First, label transfer predicted the cells as Th17.1 with high confidence (**Supplementary Fig 7B**); second, Th1- and Th17-associated genes were expressed (**Supplementary Fig 7C**); and third, Th1-polarized Th17 signature module scores were highest within these cells (**Supplementary Fig 7D**), further validating our observation.

**Figure 3:**
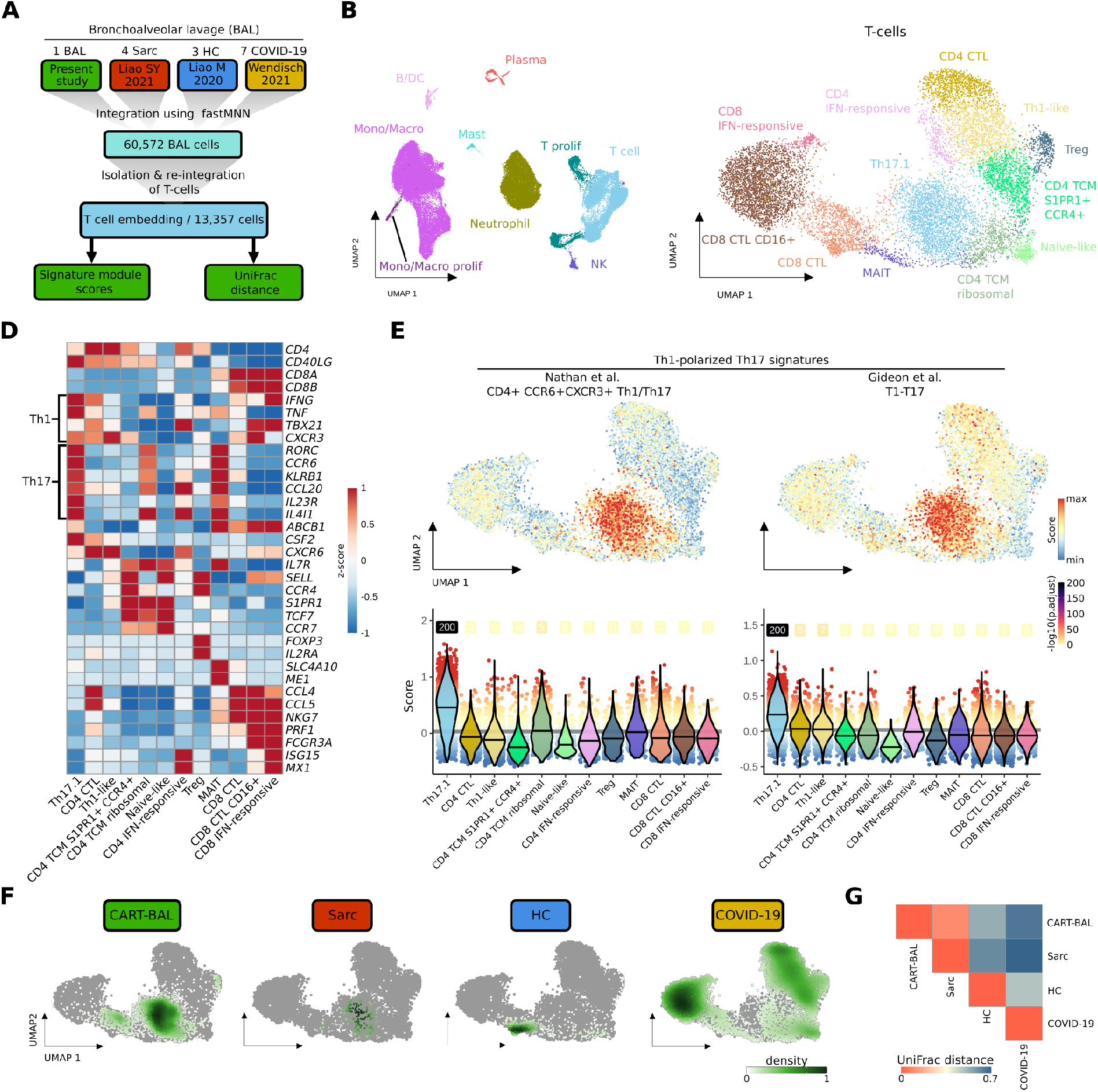
scRNA-seq data integration reveals phenotypic similarities between T-cells from CART-BAL and patients suffering from pulmonary sarcoidosis. **A**, Schematic depicting the integration workflow of BAL data from present study with BAL data from sarcoidosis, healthy controls and COVID-19 ARDS using fastMNN. T-cells were isolated and re-integrated. Signature module scores and UniFrac distances were computed. **B**, UMAP embedding of 60,572 cells from the integrated datasets colored by cell type. **C**, UMAP embedding of 13,357 T-cells isolated from the integrated datasets and re-integrated, colored by T-subsets. **D**, Heatmap showing the z-score of mean log-normalized expression of selected genes per T-subset. **E**, Cell-based gene set module scores of two Th1-polarized Th17 gene signatures. Top: Projected onto the UMAP embedding from (**c**). Bottom: depiction as violin plots across T-subsets (violin color). Lines in violins show median scores per T-subset. Grey lines indicate the average scores across all T-cells. Dot color specifies the signature module score and numbers specify -log10 transformed adjusted p-values (one-sided Wilcoxon rank-sum test against the average; -log10(p.adjust) with value ‘infinite’ (p.adjust = 0) were set to 200). **F**, Kernel density estimations of cells from the four conditions (CART-BAL, Sarc, HC, COVID-19) used in data integration shown as UMAP overlay for individual conditions. **G**, Heatmap showing the correlations between T-cells of integrated conditions by UniFrac distances. Abbreviations: BAL-bronchoalveolar lavage; Sarc - sarcoidosis; HC - healthy control; Mono/Macro - monocytes and macrophages; CTL - cytotoxic T-lymphocyte; TCM - central memory T-cell; MAIT - mucosal associated invariant T-cell.

While we observed aberrant T-cells in the CART-BAL dataset, the macrophage compartment in CART-BAL, sarcoidosis, and HC was mainly comprised of unremarkable tissue resident alveolar macrophages. A pro-fibrotic macrophage phenotype (*CD163*/*LGMN*-Mp), expressing high levels of *SPP1*, recently identified in severe COVID-19 ARDS and IPF (idiopathic pulmonary fibrosis), was absent from CART-BAL, sarcoidosis and HC BALs^26,30,32^ (**Supplementary Fig 8A-C**). The same holds true for an accumulation of neutrophils that was only present in patients with severe COVID-19 ARDS, consistent with acute infection in those patients (**Supplementary Fig 5C**).

Together, our data suggest a non-infectious, Th17.1 T-cell driven sarcoidosis-like auto-immune phenomenon located in the lungs following anti-BCMA CAR T-cell therapy. Of note, not a single plasma cell transcriptome was observed in CART-BAL, excluding resistant disease to cause the FDG signal in PET imaging.

### Discriminating myeloma relapse from sarcoidosis using PET imaging

At 6 month follow-up, the patient was still coughing. CAR T-cell numbers in the peripheral blood declined from initially 10% to 0.05% of PBMCs, and humoral immunity started to recover. FDG PET showed the known sarcoidosis-like pulmonary changes, but also revealed a number of new soft-tissue lesions located to the musculature of both lower limbs, suspicious for extramedullary relapse. In contrast to the previous scans, these lesions were CXCR4 positive (**Fig 1C, Supplementary Fig 1E**,**F**). We biopsied one of the lesions and performed scRNA-seq, revealing 99.3% of malignant plasma cells with strong *TNFRSF17* (*BCMA*) and *CCND2* expression of which a considerable amount was in a proliferative state (**Fig 4A-C**). *CXCR4* expression was detected on RNA level and expression was confirmed on protein level using flow cytometry, explaining PET positivity for CXCR4 (**Supplementary Fig 9B, Supplementary Fig 9A**). Intriguingly, a number of unexpected genes were expressed, usually seen in epithelial cells such as *EPCAM, SFN, KRT8* and *KRT18*, suggesting a plasma-epithelial transition (**Fig 4B**). EPCAM expression was also confirmed on protein level using flow cytometry (**Supplementary Fig 9A**). We compared transcriptomes from the extramedullary lesion (EMD) to two publicly available single cell datasets including one post-CAR T relapse^33^ and 13 newly diagnosed MM cases^34^. This plasma-epithelial gene expression was unique to our case (**Supplementary Fig 9B**). Whether expression of these genes was linked to extramedullary growth or even to progression from CAR T-cell therapy, has yet to be determined. The patient was later salvaged with a CXCR4-directed endo-radiotherapy (ERT), using a single intravenous injection of 90Y-labelled Pentixather (7,305 GBq), followed by high-dose melphalan and autotransplant two weeks after ERT^35^. The patient received complete remission, but relapsed 4 months after ERT and was refractory to salvage chemotherapy. Of note, respiratory symptoms completely resolved following high-dose inhalative steroids 10 weeks after autotransplant.

**Figure 4:**
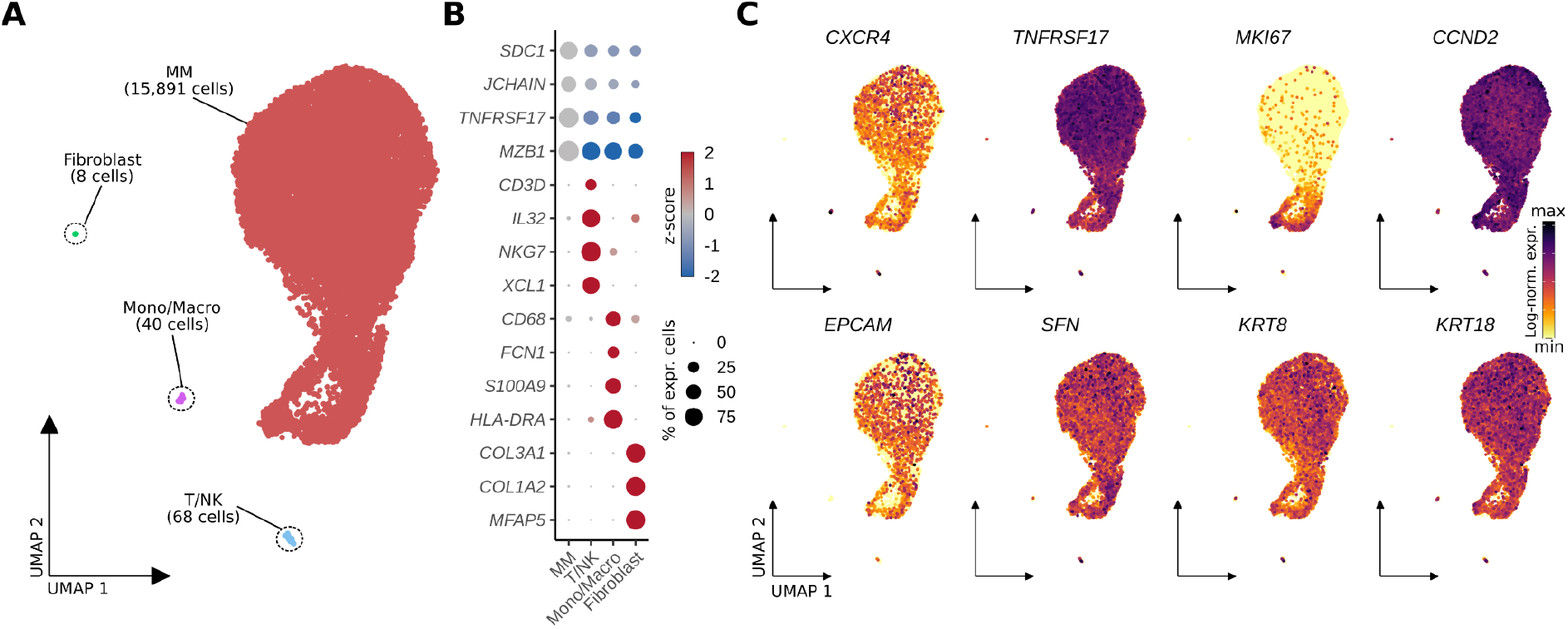
scRNA-seq analysis of a soft-tissue lesion in the lower limb six months after anti-BCMA CAR T-cell therapy showed expression of epithelial-associated genes in multiple myeloma cells. **A**, UMAP embedding of 16,007 single cells from an extramedullary soft-tissue lesion colored by cell type. **B**, Dotplot showing scaled expression (color) of respective marker genes per cell type. Dot size depicts the percentage of non-zero expressing cells. **C**, Log-normalized gene expression of *CXCR4*, the CAR T target antigen *TNFRSF17* (*BCMA*), proliferation marker *MKI67, CCND2* (marking malignancy), as well as epithelial cell associated genes (*EPCAM, SFN, KRT8, KRT18*) color-coded and projected onto the UMAP embedding from (**A**). Abbreviations: Mono/Macro - monocytes/macrophages.

## DISCUSSION

We hypothesize that CRS after CAR T-cell infusion triggered expansion of pathogenic Th17.1 T-cells in the lung leading to sarcoidosis-like manifestations. Th1-polarized Th17 cells are key effectors of autoimmune inflammation^12^ including inflammatory bowel disease^13^, multiple sclerosis^36^, and sarcoidosis^17^. T-cells with Th17.1 characteristics have been described to be enriched in the bone marrow of MM patients^37^. Of note, our patient suffered from rare pulmonary EMD at the time of diagnosis. Thus, preexistence of Th17.1 T-cells in the lung prior to CAR T-cell treatment is likely and could represent a risk factor for sarcoidosis-like reactions. Unmasking of autoimmune diseases is a known side effect after checkpoint inhibitors^38,39^, and here we describe a similar phenomenon after CAR T therapy.

PET imaging using multiple tracers is a noninvasive approach to decode tumor heterogeneity in MM^40^. Using the three tracers FDG, DOTATOC, and CXCR4-targeted Pentixafor, we were able to discriminate between autoimmune inflammation and MM relapse. In light of the plethora of novel tracers currently entering nuclear medicine^40,41^, PET imaging beyond FDG is a promising tool in the follow-up assessment of CAR T treated patients allowing for discrimination between inflammation and true relapse. Of note, in the absence of known pulmonary MM manifestations, sarcoidosis on FDG PET/CT is a known pitfall, with typical imaging features on PET, and usually does not need confirmation using alternative radiotracers. However, our case study is an illustrative example on how multi tracer PET imaging could help to narrow down the differential diagnosis. Another potential biomarker helping to distinguish MM progression from immune-mediated changes is soluble (s)BCMA. A limitation, however, could be that sBCMA clearance takes several months and flare up phenomena typically occur early after initiation of immunotherapy. Moreover, we and others have reported on MM progression after anti-BCMA CAR T-cell therapy lacking sBCMA increase due to clonal biallelic *BCMA*-loss^33,42^.

Finally, we report the case of a BCMA positive relapse after anti-BCMA CAR T. While antigen loss is a rare tumor-intrinsic mechanism of resistance^33,42^, the biology underlying relapse with preserved antigen expression is poorly understood. In our case, relapse occurred along with recovery of non-involved immunoglobulins, suggesting loss of CAR T-cell function and subsequent outgrowth of residual MM cells. Larger trials are warranted to investigate whether the plasma-epithelial signature that we described contributed to relapse. Appreciating the lack of activity of CAR T cells in carcinoma and the limited duration of response to CAR T in EMD compared to patients without EMD^43^, plasma-epithelial transformation could alter CAR T-cell activity and confer a mechanism of resistance.

In conclusion, we demonstrated a clinically relevant autoimmune phenomenon after CAR T-cell therapy that can be delineated using scRNA-seq and PET imaging.

## METHODS

### Ethics

The patient gave written informed consent for scientific evaluations. The study was approved by the internal review board of the University of Würzburg (reference # 8/21) and adhered to the tenets of the Declaration of Helsinki of 2008.

### Radiotracer Synthesis, Image Protocols and Interpretation

We used a fully automated synthesis module (Scintomics, Fürstenfeldbruck, Germany) to synthesize ^68^Ga-Pentixafor, as described previously^35^. ^18^F-FDG and ^68^Ga-DOTATOC were prepared as described previously^44,45^. PET/CTs were carried out using a dedicated PET/CT scanner (Siemens Biograph mCT 64 or mCT 128; Siemens Medical Solutions, Germany). Prior to administration of ^18^F-FDG, patients fasted for at least 6 h with blood glucose levels below 160 mg/dl at time of scan. For ^68^Ga-Pentixafor and ^68^Ga-DOTATOC PET/CT, fasting was not required. Spiral CT with or without intravenous contrast was conducted and included a field of view from the base of the skull to the proximal thighs. PET emission data were acquired in three-dimensional mode (200 × 200 matrix, 2–3 min emission time per bed position). After decay and scatter correction, PET data were then reconstructed iteratively using the algorithm provided by Siemens Esoft (Siemens, Erlangen, Germany). Image interpretation was conducted on a visual basis and carried out by an expert reader (RAW).

### Sample preparation and scRNA-seq processing

CART-BAL sample was obtained at 3-month follow-up, diluted in PBS and filtered through a 40 μm mesh resulting in a single cellm mesh resulting in a single cell suspension. EMD sample was obtained at 6-month follow-up by ultrasound guided push biopsy. The biopsy was filtered with a 100 μm mesh resulting in a single cellm strainer followed by tissue digestion using Collagenase A (10 mg/mL), Collagenase D (10 mg/mL), DNAse I (2 mg/mL). Digestion was stopped with EDTA (100 mM) followed by a second filtering through a 70 μm mesh resulting in a single cellm mesh resulting in a single cell suspension.

Obtained single cell suspensions were adjusted to a concentration of 400 cells/μm mesh resulting in a single celll and 1000 cells/μm mesh resulting in a single celll for CART-BAL and EMD, respectively, and loaded into the 10x Chromium controller for scRNA-seq. Following the detailed protocol provided by 10x genomics, single cell reagent kit v3.1 was used for reverse transcription, cDNA amplification and library construction. Libraries were quantified using QubitTM 2.0 Fluorometer (ThermoFischer) and quality was checked by 2100 Bioanalyzer with High Sensitivity DNA kit (Agilent). S2 flow cells were used for sequencing on a NovaSeq 6000 sequencer (Illumina). CART-BAL has been run in duplicate to obtain technical replicates.

### Single-Cell RNA-seq analysis

#### scRNA-seq analysis of CART-BAL dataset

Raw sequencing data was demultiplexed and quality-checked using the CellRanger (v.3.1.0) ‘mkfastq’ script. Alignment and transcript quantification was performed using the CellRanger ‘count’ script against the GRCh38 (Ensembl93) human genome assembly. Ambient RNA signals were removed using SoupX to mitigate potential effects of RNA released from dead or ruptured cells^46^. The obtained SoupX-adjusted count matrices were loaded into R (4.1.0) and downstream analysis was performed using the Seurat R package (v4.1.1)^47^. Low quality transcriptomes, as well as epithelial cells (based on expression of *EPCAM*) were removed. Quality filtering thresholds for the number of detected genes, number of detected unique RNA molecules (UMIs) and fraction of mitochondrial UMI-counts are available in **Supplementary Table 1**. Count data was log-normalized (Seurat function NormalizeData), highly variable features were identified (Seurat function FindVariabeFeatures, n.features = 5000) and scaled (Seurat function ScaleData). Principal component analysis (PCA) was calculated using RunPCA function of Seurat based on highly variable genes. A two-dimensional embedding was computed with the uniform manifold approximation and projection (UMAP) algorithm ^48^ using the first 45 PCA-dimensions (Seurat function RunUMAP). Nearest neighbor graph construction was performed using the Seurat function FindNeighbors (dims = 1:45). Unsupervised clustering was applied using the Louvain algorithm implemented in Seurat (Seurat function FindClusters) with the resolution parameter set to 1.8.

For analysis of CART-BAL T-cells respective clusters (T CD4, TCD8, Treg) were isolated from the dataset using the subset function and the subset was reanalysed repeating basic Seurat steps (described above) based on 1000 highly variable features, the first 15 PCA-dimensions, and clustering resolution parameter set to 0.7.

#### Analysis of integrated BAL datasets

BAL Single cell transcriptomes generated in this study (CART-BAL) and three previous studies, including BAL samples from four pulmonary sarcoidosis patients^27^, three healthy donors^25^ and seven patients with severe COVID-19 acute respiratory distress syndrome (ARDS) with profound fibrotic remodeling^26^, were integrated into a single embedding. The integration workflow is sketched in **Supplementary Fig 5**. Raw sequencing data were acquired from Wendisch *et al*.^26^ (COVID-19) and Liao M. *et al*.^25^ (HC). Alignment was performed using a common CellRanger version (v3.1.0) and a common reference (GRCh38, Ensembl93) to avoid confounding factors from this step, as the recently pre-printed human lung cell atlas has demonstrated the CellRanger version to constitute a key factor of variance, especially in the T-cell lineage^49^. After obtaining count-matrices, SoupX was used to remove ambient RNA signals in each dataset^46^. As Sarcoidosis BAL scRNA-seq data was not available as raw sequencing data due to patient privacy restrictions, count level data were acquired from this study (Sarc). These data were aligned with CellRanger v3.1.0 to reference GRCh38 Ensembl93 in the original study^27^. The datasets were further processed, and quality control was performed using Seurat v4.1.1 based on customized thresholds for the number of detected genes, number of detected unique RNA molecules (UMIs) and fraction of mitochondrial UMI-counts (**Supplementary Table 1**). In addition, epithelial and erythrocyte clusters were removed. All datasets were merged, and initial basic Seurat analysis was performed using 3000 highly variable genes and 55 first PCA-dimensions. As confounding covariate “patient” for the CART-BAL datasets (two replicates/datasets, one patient), COVID-19 datasets (7 datasets, 7 patients), and sarcoidosis datasets (4 datasets, 4 patients) as well as “donor” for healthy controls (3 datasets, 3 donors) were identified (**Supplementary Fig 5B**). Data integration correcting for these covariates was performed using the fastMNN implementation of the Bioconductor batchelor package based on the 3000 highly variable genes^50^. The fastMNN approach for data integration was chosen as it was one of the top-performers in a recent benchmark by Luecken et al., especially in the human immune cell task^51^. For computation of the UMAP embedding and nearest neighbor graph construction 55 first MNN-dimensions were used. Unsupervised clustering, based on the nearest neighbor graph, was performed using the Louvain algorithm with the resolution parameter set to 0.4.

Of note, dataset HC4 from the Liao M. et al. paper was excluded due to patient privacy restrictions. In the original study alignment was performed using a different CellRanger version and reference as desired for the integration pipeline in the present study^32^.

T-cells and monocyte/macrophage (Mono/Macro) cells were separated from the integrated dataset and reanalysed, repeating integration as described above. For T-cells and monocyte/macrophage subsets 3000 and 1000 highly variable genes, 55 and 25 first PCA-dimensions, 55 and 25 first MNN-dimensions, and clustering resolutions of 1.5 and 0.4 were used, respectively, for dimensional reduction and Louvain clustering.

#### Projection of T-cells from cutaneous sarcoidosis to integrated BAL T-cell embedding

T-cells of a publicly available dataset including three patients suffering from cutaneous sarcoidosis were extracted after preprocessing using the common CellRanger version (v3.1.0) and reference (GRCh38, Ensembl93) as well as quality filtering^31^ (**Supplementary Table 1**). Seurat v4.1.1 was used for mapping of obtained T-cell transcriptomes following the steps detailed in the author’s tutorial^47^. Mapping anchors were identified using the function FindIntegrationAnchors with k.filter = 100, dims = 1:10, and reference.reduction = “pca’’ (as recommended for unimodal analysis). MapQuery function was run using the identified mapping anchors with reference.reduction = “pca” and T-cell subsets (from **Fig 3C**) as refdata for label transfer. Query cells with a label transfer prediction score below 0.4 were labeled as unassigned.

#### Analysis of EMD sample

Raw sequencing data was handled in the same way as described above. Quality filtering thresholds for the number of detected genes, number of detected unique RNA molecules (UMIs) and fraction of mitochondrial UMI-counts are available (**Supplementary Table 1**). basic Seurat steps (described above) were based on 1000 highly variable genes and 10 first PCA-dimensions. Seurat function CellSelector was used to select MM cell and non-MM cell clusters based on UMAP-coordinates, as unsupervised clustering could not resolve non-MM cell clusters within reasonable clustering resolutions due to their low cell numbers. The described expression of epithelial-associated genes in MM cells was discovered during exploratory data analysis. For gene expression comparison between the EMD sample and intramedullary (IMD) MM samples additional publicly available datasets including one post-CAR T MM case^33^ and 13 newly diagnosed MM cases^34^ were acquired, quality filtered (**Supplementary Table 1**), and plasma/MM cells were extracted. Count data was log-normalized. Differential expression analysis results between EMD and IMD can be found in **Supplementary Table 2**.

#### Annotation of major cell types and subpopulations

Major cell types were annotated based on expression of canonical marker genes (AMp: *CD68, CD163, FABP4, MARCO*; monocytes: *VCAN, FCN1*; mDC: *CD1C, CD1E, FLT3, LGALS2*; cDC1: *FLT3, LGALS2, CLEC9A, XCR1*; neutrophils: *FCGR3B, CXCL8, CSF3R*; *G0S2*; Mast cells: *TPSAB1, TPSB2, MS4A2, GATA2*; T-cells: *CD3D, CD3E, IL32*; CD4 T-cells: *CD4*; CD8 T-cells: *CD8A, CD8B*; Tregs: *FOXP3, IL2RA*; NK-cells: *TRDC, NKG7, KLRC1, XCL1, XCL2, KLRF1;* B-cells: *MS4A1, SPIB*; Plasma-cells: *JCHAIN, SDC1, MZB1, TNFRSF17, XBP1*; Fibroblasts: *COL3A1, COL2A1, MFAP5;* proliferating cells: *MKI67, TOP2A*). T-cell subpopulations were annotated upon careful assessment of expressed genes. particularly, expression of canonical lineage and cell state markers was interrogated (Th1: *IFNG, TNF, TBX21, CXCR3*; Th17: *IL17A, CCL20, RORC, CCR6, KLRB1*; central memory T-cell (TCM): *CCR7, TCF7, S1PR1, IL7R, SELL*; naive T-cells: *CCR7, TCF7, LEF1*; MAIT: *SLC4A10, ME1*; cytotoxic lymphocytes (CTL): Granzymes, *PRF1*; CD4 CTL: *CD4, CCL4, CCL5, PRF1*; IFN-responsive: *ISG15, MX1, RSAD2*). Macrophage subpopulations were annotated in an analogous fashion (CD163/LGMN Mp: *SPP1, LGMN*; pro-inflammatory AMp: *CCL3, CCL4, CCL20, TNFAIP6, SOD2*; metallothionein AMp: metallothioneins, e.g. *MT1G, MT2A, MT1X*; IFN-responsive: *ISG15, MX1, RSAD2*; Foam cells: *LDLR, SQLE, HMGCR, MSMO1*).

#### UniFrac distance analysis

To test if T-cells from different conditions in the integrated BAL analysis tended to form the same unsupervised clusters, UniFrac distance analysis was applied^30^. Mean principal component (PC) values were calculated across significant PCs for each unsupervised cluster (see details above). Euclidean distance was calculated for each PC between clusters and a distance matrix was built with decreasing weights for PCs, analogous to decreasing eigenvalues of PCs. The distance matrix was used as input for hierarchical clustering with the complete linkage method (R function hclust) to obtain a precalculated tree. Code from the scUniFrac R package was modified to enable use of this precalculated tree for computation of generalized UniFrac distances between the conditions^52,53^ (code available on GitHub).

#### Signature module score calculation

Single cell level scores for signatures were computed using the Seurat function AddModuleScore. In brief, genes were binned (nbin = 24) based on their average expression in all cells. Expression of genes from a given signature was calculated and subtracted by the aggregated expression of a control feature set (ctrl = 100) selected randomly from the bins the analysed signature genes were part of. Statistical significance was assessed by pairwise, one-sided (alternative = “greater”) Wilcoxon rank-sum test (R function wilcox.test) of each group compared to the average. Signatures were retrieved from the literature. Signatures and respective sources are listed in **Supplementary table 1**.

#### Differential expression analysis

Differential expression analysis was performed using the Wilcoxon rank-sum test implementation of the Seurat function FindAllMarkers (only.pos = TRUE). Differential expression analysis results are available in **Supplementary Table 2**.

### Flow cytometry

EMD cells were thawed in RPMI supplemented with 10% FCS. Staining was performed in PBS at 4 °C with the following antibodies: CD184/CXCR4 – PerCP/eFluor710 (1:100, 12G5, Thermofisher), CD326/EpCAM – APC (1:100, 9C4, Biolegend), CD269/BCMA – PE (1:100, REA361, Miltenyi), CD38-FITC (1:100, HIT2, Thermofisher), CD138 – BV510 (1:20, MI15, Biolegend), CD45 – BV605 (1:20, 2D1, Biolegend), CD3 – AF700 (1:100, HIT3a, Biolegend). For dead cell exclusion, Fixable Viability Dye eFluor ™ 780 (1:1000, Thermofisher) was used. A FcGr blocking step was not performed. Events were acquired on the Attune NxT (Thermofisher). Data was analyzed with FlowJo version 10.8.0.

### Statistical analysis

The non-parametric two-sided Wilcoxon rank-sum test was used to compare gene expression values between groups of cells. A p-value < 0.05 was considered as statistically significant.

### Code availability

All R code used for scRNA-seq data analysis has been deposited on GitHub: https://github.com/alexleipold/SarcoidosisAfterCART

### Data availability

Data generated in the present study are available from the authors upon reasonable request. Healthy BAL datasets from Liao M. et al.^25^ are accessible from the Gene Expression Omnibus (GEO) database under accession no. GSE145926. COVID-19 ARDS BAL data from Wendisch et al.^26^ are available at the European Genome-Phenome Archive (EGA) under study ID EGAS00001004928. Sarcoidosis BAL data from Liao S.Y. et al.^27^ is accessible on GEO under accession no. GSE184735. Data from Damsky et al.^31^, including three patients with cutaneous sarcoidosis, is available on GEO with accession no. GSE169147. The dataset of a post-CAR T relapsed multiple myeloma patient from Da Vià et al.^33^ can be accessed on GEO with accession no. GSE143317. Newly diagnosed MM plasma cell datasets from de Jong et al.^34^ are available on ArrayExpress, no. E-MTAB-9139.

## Data Availability

All data produced in the present study are available upon reasonable request to the authors.

https://www.ncbi.nlm.nih.gov/geo/query/acc.cgi?acc=GSE145926

https://ega-archive.org/studies/EGAS00001004928

https://www.ncbi.nlm.nih.gov/geo/query/acc.cgi?acc=GSE184735

https://www.ncbi.nlm.nih.gov/geo/query/acc.cgi?acc=GSE169147

https://www.ncbi.nlm.nih.gov/geo/query/acc.cgi?acc=GSE143317

https://www.ebi.ac.uk/biostudies/arrayexpress/studies/E-MTAB-9139

---

**Supplementary Figure 1:**
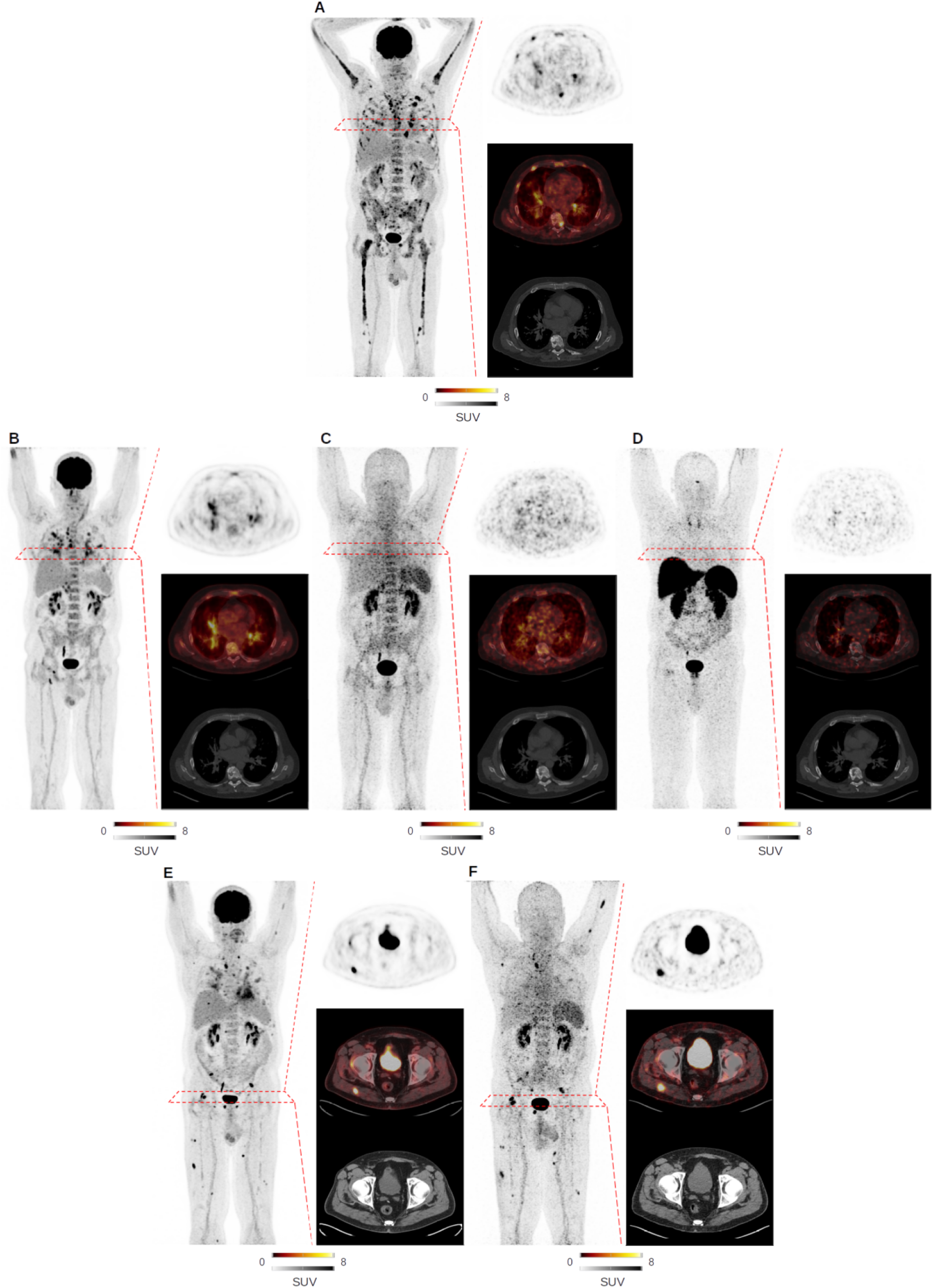
Whole body PET imaging at baseline and during follow-up using multiple PET tracers along with maximum intensity projection (MIP) on the right and transaxial PET, PET/CT and CT on the left. **A**, FDG-PET at baseline shows multiple focal lesions at the axial and appendicular skeleton. Respective transaxial slides revealed multiple disease manifestations in the ribs. **B-D**, Follow-up assessment 3 months after CAR T infusion shows full resolution of focal lesions, but residual FDG uptake located to the lung and mediastinal lymph nodes, as visualized on the transaxial slides. Matched simultaneous CXCR4-targeted Pentixafor (**C**) and SSTR-directed DOTATOC (**D**) scans did not show any focal uptake. Respective transaxial slides on the identical site as chosen for FDG scan are shown. **E**,**F**, Matched simultaneous FDG (**E**) and CXCR4-directed Pentixafor (**F**) PET 6 months after CAR T infusion depicted new focal lesions in line with relapse, as seen on transaxial slides at the pelvis.

**Supplementary Figure 2:**
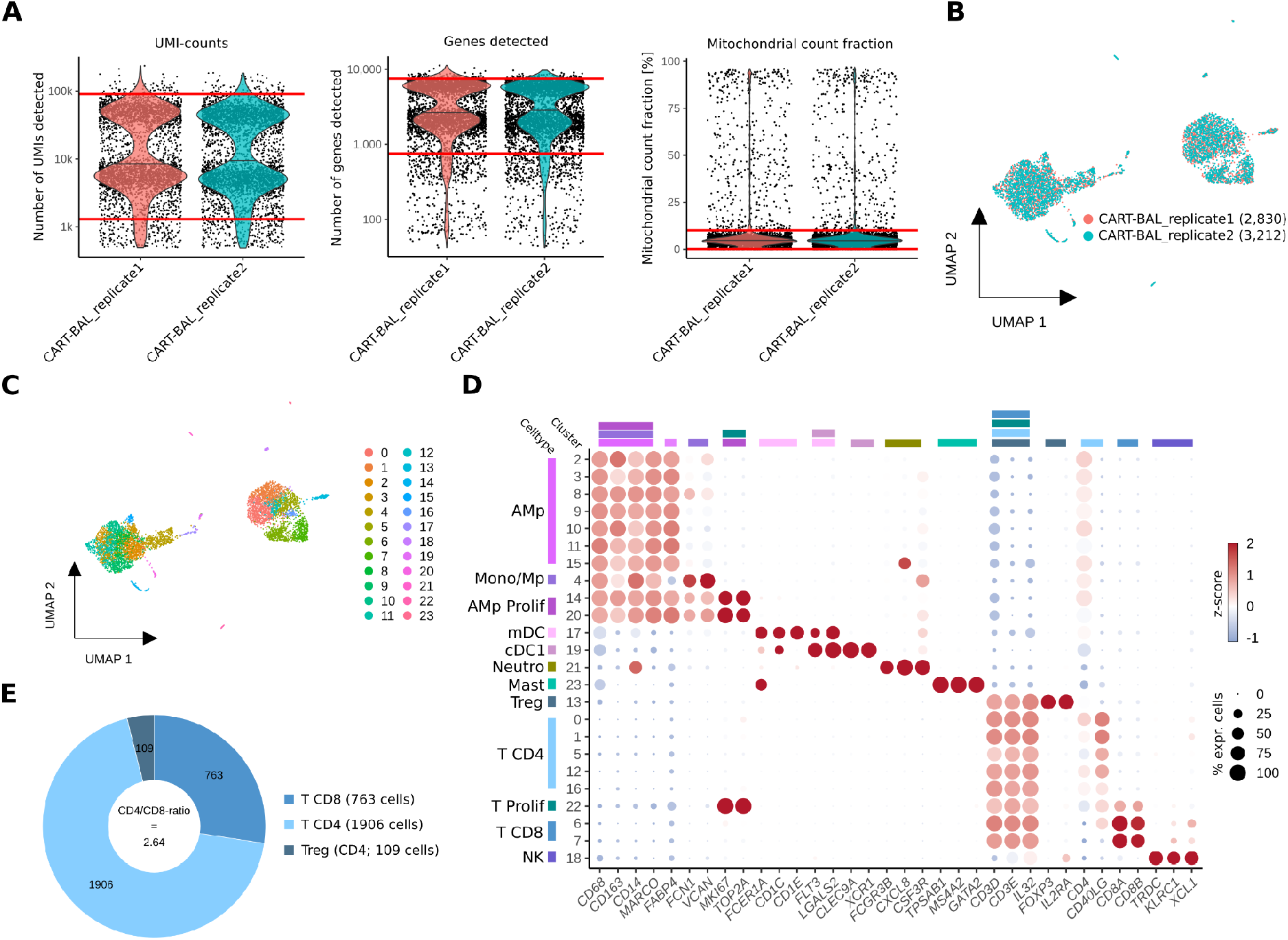
scRNA-seq analysis of BAL from a MM patient at 3-months follow-up of a CAR T therapy (CART-BAL) in technical duplicates. **A**, Data quality metrics and filtering across two scRNA-seq CART-BAL technical duplicates depicted in violin plots (replicate1: 3,764 cells, replicate2: 4,180 cells). Red lines indicate the thresholds that were used for quality control filtering. For UMI-counts and genes detected, log10-scale was used. **B**, UMAP embedding visualising scRNA-seq of 6,042 cells from CART-BAL color-coded by technical replicate. Numbers in brackets indicate cell number per replicate after quality control filtering (panel a). **C**, UMAP embedding as in (**B**) partitioned in clusters and color-coded by Louvain cluster. **D**, Dotplot showing log-normalized, scaled expression (color) and the proportion in percentage of non-zero expressing cells (size) of canonical marker genes per cluster identified in the UMAP (panel c). **E**, Pie plot depicting the proportions of CD4 cells, Tregs, and CD8 cells within T-cells, as well as the CD4/CD8-ratio. Abbreviations: AMp - alveolar macrophage; Mono/Mp - monocyte/macrophage; Neutro - neutrophil; Prolif: Proliferating.

**Supplementary Figure 3:**
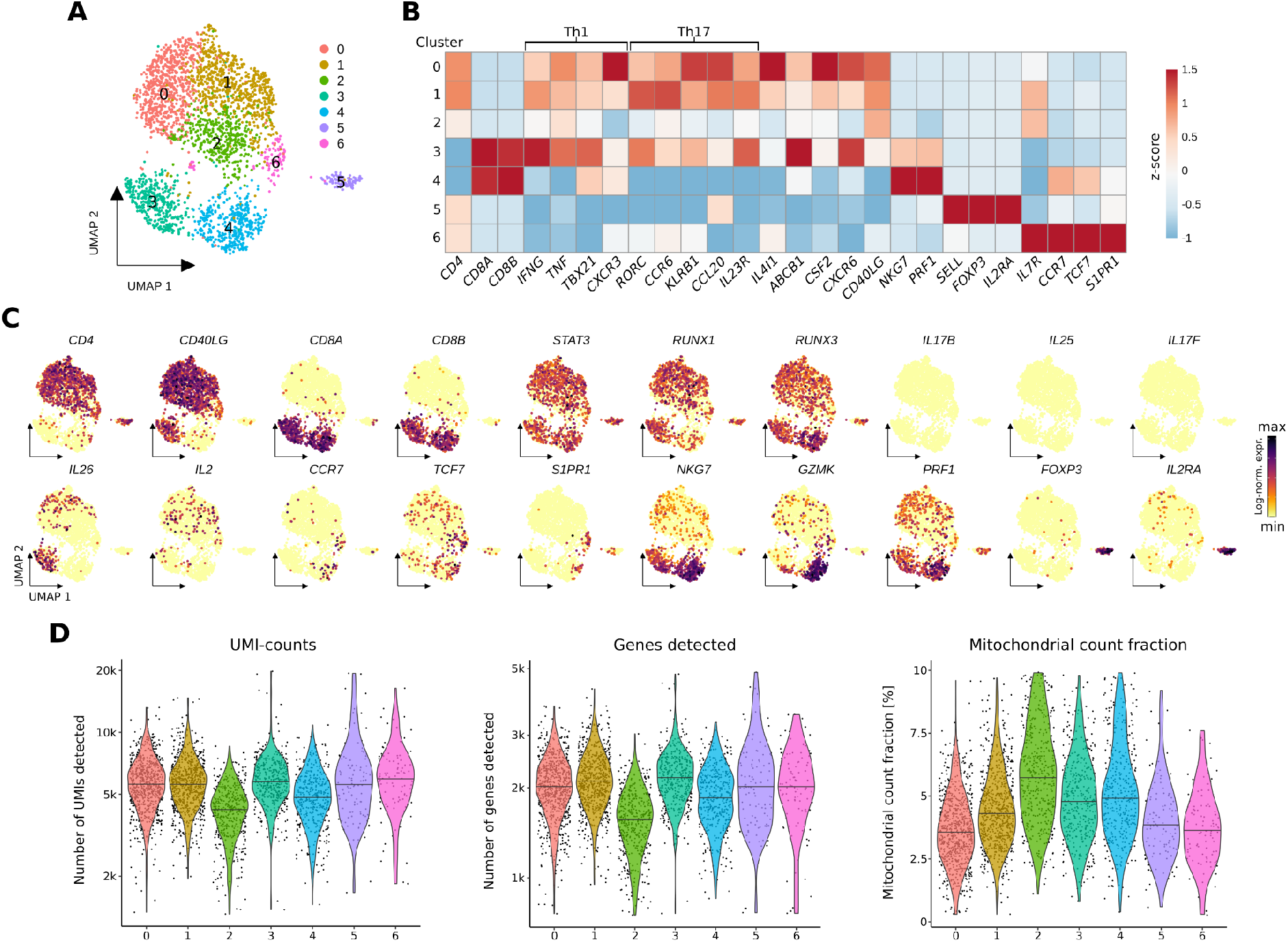
scRNA-seq analysis of the T-cell compartment of the CART-BAL from a MM patient at 3-months follow up after anti-BCMA CAR T-cell therapy in technical duplicate. **A**, UMAP embedding of single-cell transcriptomes of T-cells from the CART-BAL colored and numbered by Louvain cluster. **B**, Heatmap showing the z-score of mean log-normalized expression of selected genes per cluster identified in panel a. Th1- and Th17-associated genes are indicated by brackets. **C**, Log-normalized gene expression of selected T-cell markers color-coded and projected on the UMAP embeddings from (**A**). **D**, Violin plots depicting UMI-counts, number of genes detected and percentage of mitochondrial genes across the T-cell clusters identified in (**A**).

**Supplementary Figure 4:**
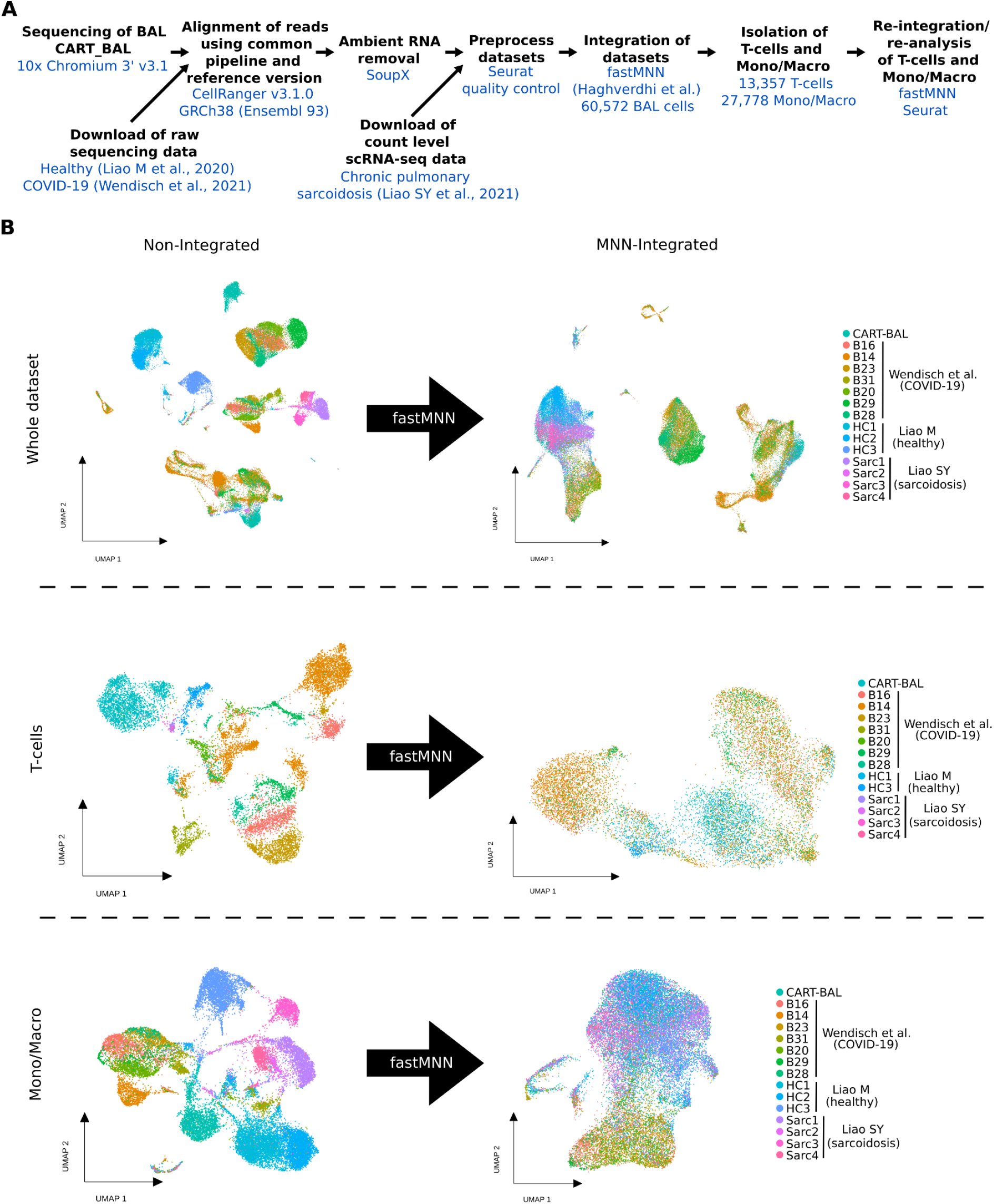
Dataset integration. **A**, Schematic of the workflow used for data integration. **B**, UMAP embeddings of the merged BAL datasets with and without MNN-integration colored by patient or donor for the whole dataset (60,572 cells), T-cells (13,357 cells) and monocyte/macrophages (Mono/Macro; 27,778 cells). Abbreviations: BAL - bronchoalveolar lavage.

**Supplementary Figure 5:**
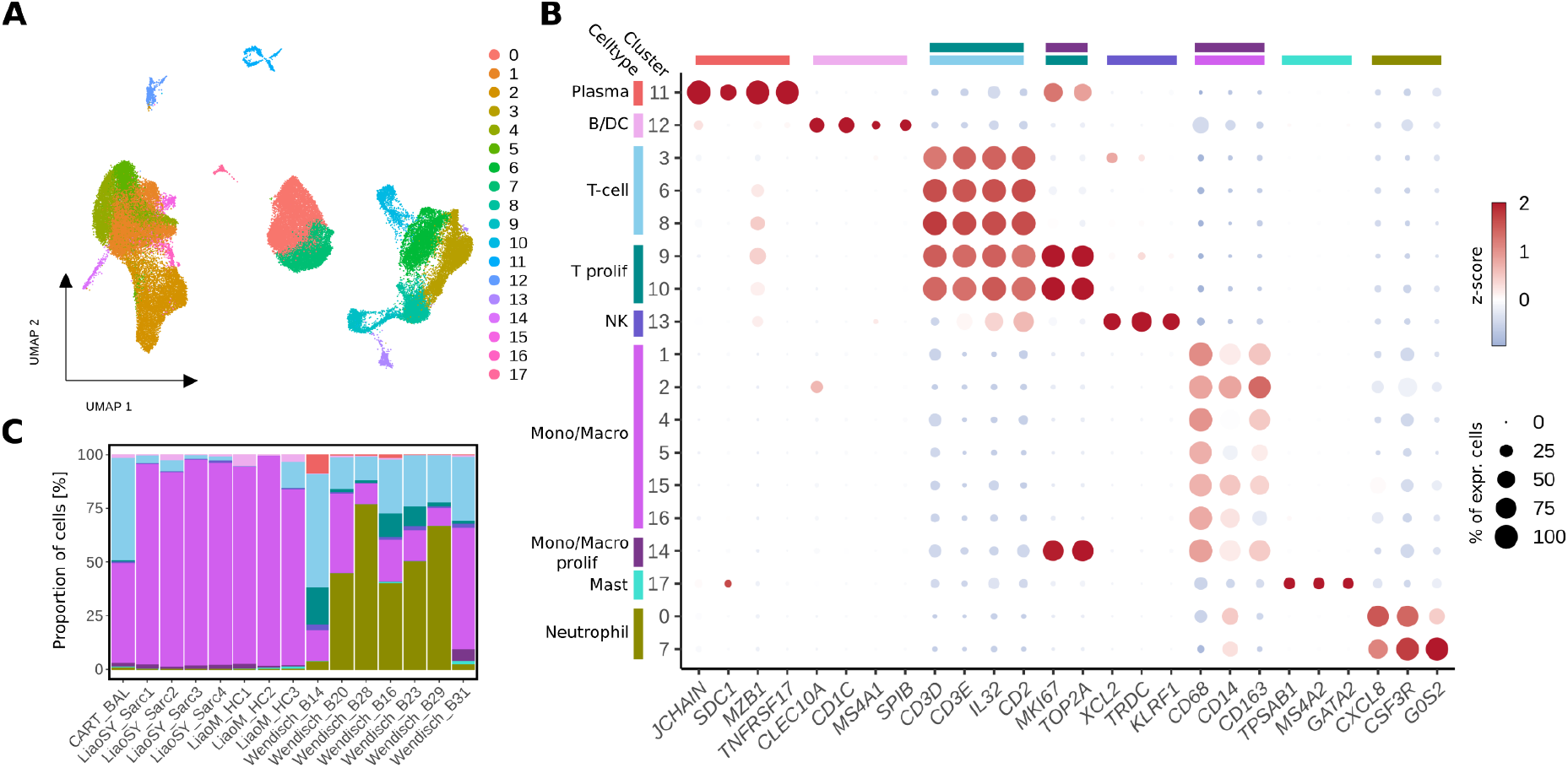
Analysis of integrated scRNA-seq BAL datasets from CART-BAL, Sarcoidosis, healthy control, COVID-19. **A**, UMAP embedding of integrated BAL cells obtained after data integration (**Supplementary Fig 4B**) colored by the Louvain cluster. **B**, Dotplot showing log-normalized, scaled expression (color) and the proportion in percentage of non-zero expressing cells (circle size) of canonical marker genes per cluster and annotated cell types. **C**, Relative proportions of cell types across patients and donors (Liao S.Y. et al. - sarcoidosis; Liao M. et al. - healthy control; Wendisch et al. - Covid-19). Cell Types are color coded as in (**B**). Abbreviations: Mono/Macro - monocyte/macrophage; prolif – proliferating.

**Supplementary Figure 6:**
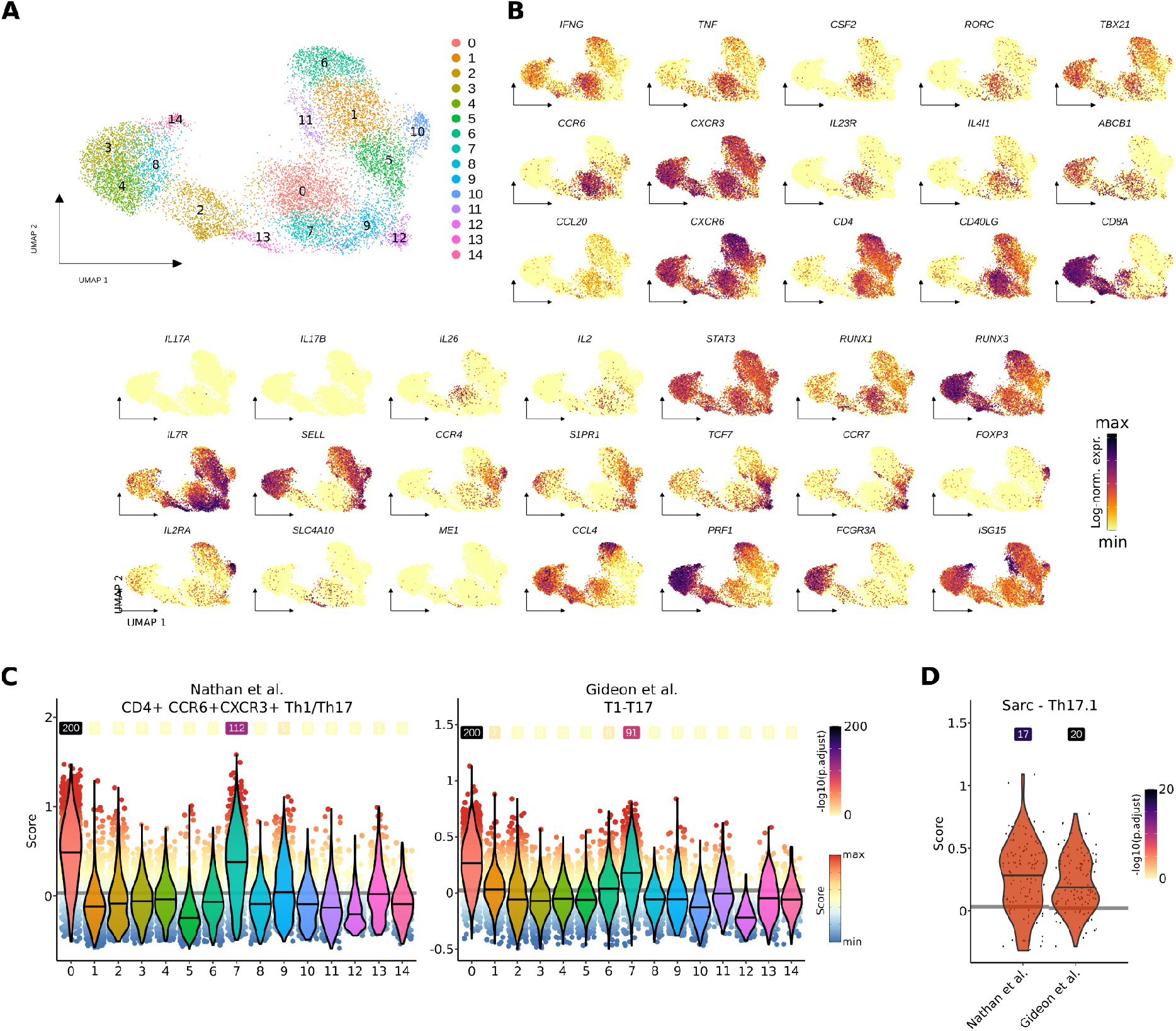
scRNA-seq analysis of T-cells isolated from the integrated BAL datasets. **A**, UMAP embedding of T-cells (T-cells, **Fig 3B and Supplementary Fig 4B**) from integrated BAL datasets of four conditions colored by Louvain clusters. **B**, Gene expression level of selected genes log-normalized, color-coded and projected on the UMAP embeddings of T-cells from integrated BAL datasets. **C**, Cell-based gene set module scores of two Th1-polarized Th17 gene signatures depicted as violin plots across Louvain clusters. Lines in violins show median scores per cluster. Grey lines indicate the average scores across all T-cells. Dot color specifies the signature module score and numbers specify -log10 transformed adjusted p-values (one-sided Wilcoxon rank-sum test against the average; -log10(p.adjust) with value ‘infinite’ (p.adjust = 0) were set to 200). **D**, Scores from (**C**) for Th17.1 cells (see **Fig 3C**) from the condition sarcoidosis (Sarc). Lines in violins show median scores per signature. Grey lines indicate the average scores across all T-cells for the respective signature. Numbers specify -log10 transformed adjusted p-values (one-sided Wilcoxon rank-sum test against the average of all T-cells). Abbreviations: TCM - central memory T-cell; CTL - cytotoxic T-lymphocyte.

**Supplementary Figure 7:**
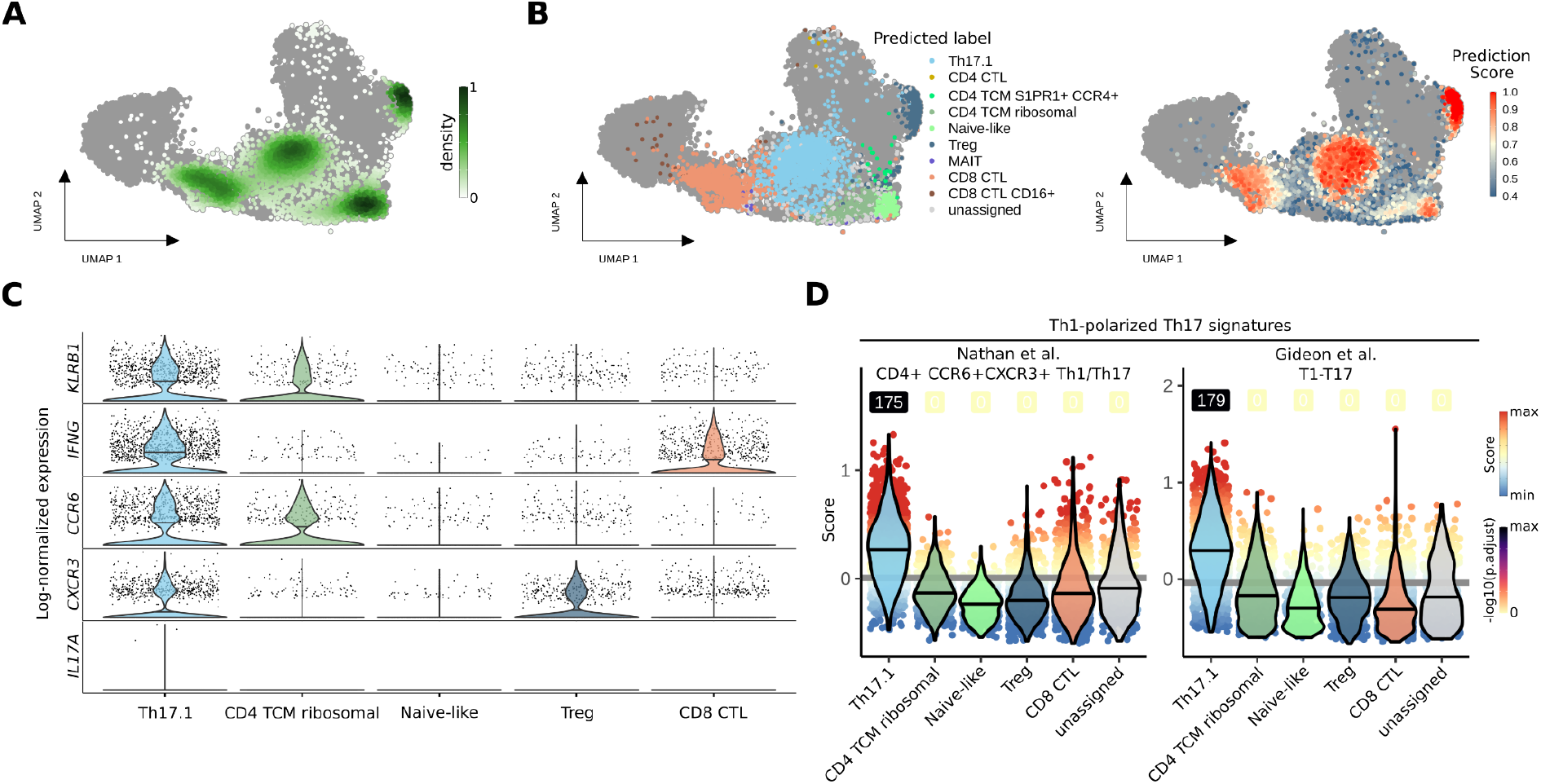
Projection of single-T-cell transcriptomes from skin sarcoidosis biopsies (Damsky *et al*, 2022) onto the integrated BAL T-cell embedding. **A**, Overlay of kernel density estimation of the skin sarcoidosis T-cells on the integrated BAL T-cell UMAP embedding. **B**, Label transfer predictions (left) and prediction scores (right) as overlay on the integrated BAL T-cell UMAP embedding. **C**, Depiction of log-normalized expression of selected genes in skin sarcoidosis T-cells as violin plots across predicted T-cell subsets. Predicted T-cell subsets with more than 30 assigned cells are shown. **D**, Cell-based gene set module scores of two Th1-polarized Th17 gene signatures in skin sarcoidosis T-cells depicted as violin plots across predicted T-cell subsets. Lines in violins show median scores per predicted T-cell subset. Grey lines indicate the average scores across all projected T-cells. Dot color specifies the signature module score and numbers specify-log10 transformed adjusted p-values (one-sided Wilcoxon rank-sum test against the average). Predicted T-cell subsets with more than 30 assigned cells and unassigned cells are shown. Abbreviations: CTL - cytotoxic T-lymphocyte; TCM - central memory T-cell; MAIT - mucosal associated invariant T-cell.

**Supplementary Figure 8:**
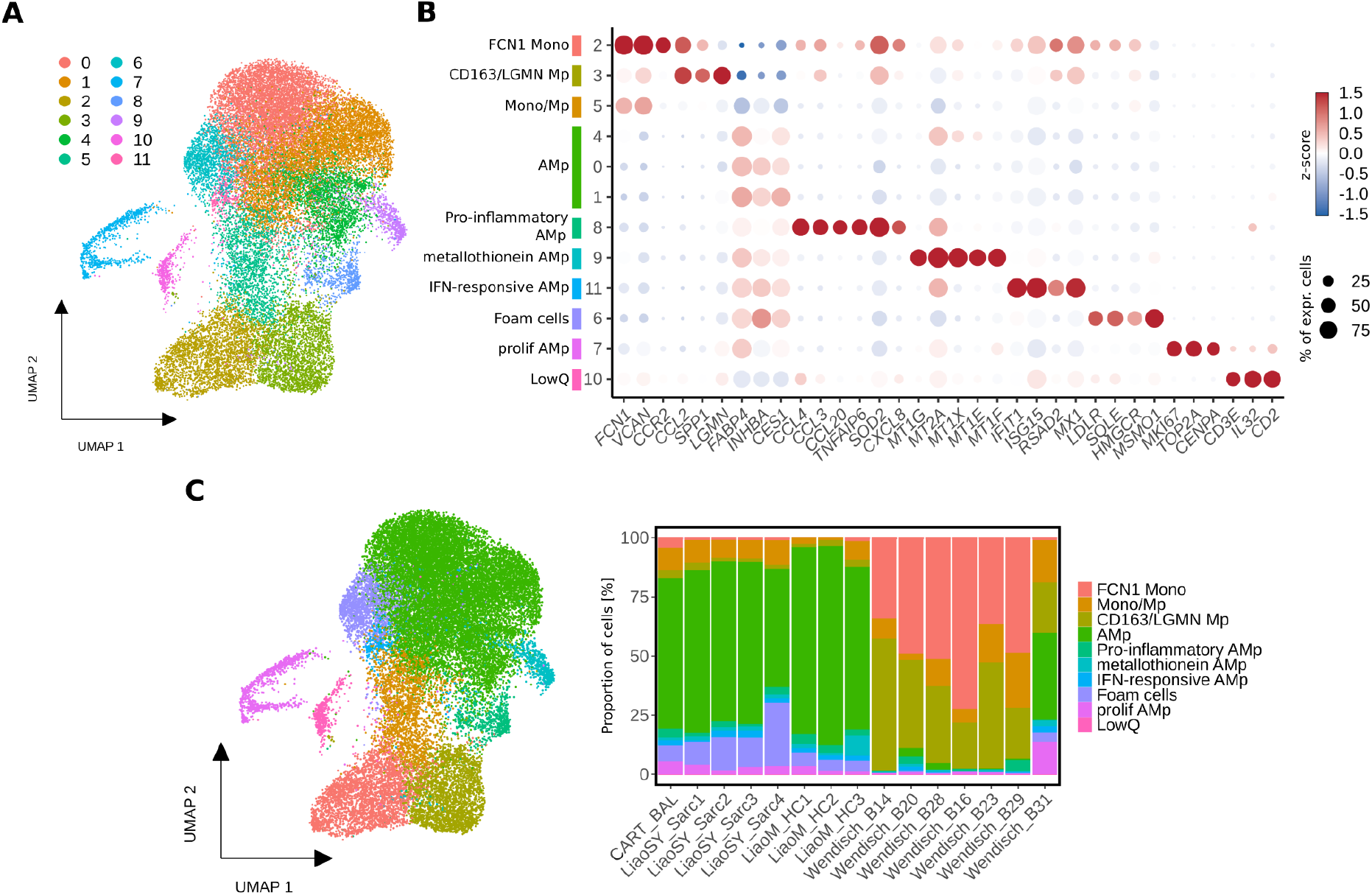
scRNA-seq analysis of monocytes/macrophages isolated from the integrated BAL datasets. **A**, UMAP embedding of 27,778 monocytes/macrophages (Mono/Macro, **Fig 3B and Supplementary Fig 4B**) from integrated BAL datasets of four conditions colored by Louvain clusters. **B**, Dotplot showing log-normalized, scaled expression (color) and the proportion in percentage of non-zero expressing cells (size) of canonical marker genes per cluster. Respective subset annotation with color code is shown as colored bars (left). **C**, Mono/Macro subset annotation plotted as UMAP embedding and proportion bar plot over individual patients/donors. Abbreviations: Mono - monocyte; Mp - macrophage; AMp - alveolar macrophage; LowQ - low quality cell.

**Supplementary Figure 9:**
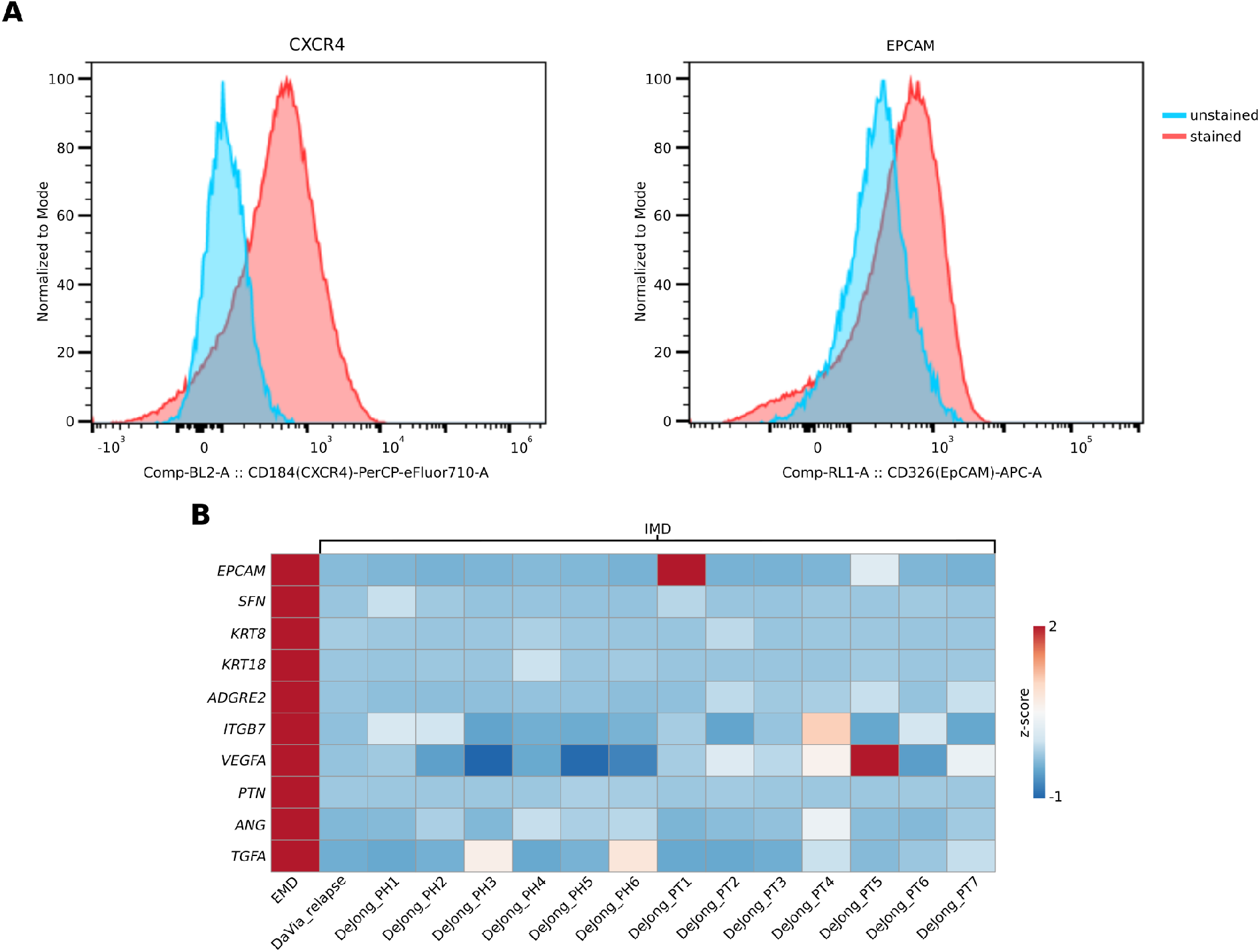
FACS analysis of EMD sample and comparison of EMD sample scRNA-seq expression of selected genes to publicly available IMD scRNA-seq samples. **A**, CXCR4 and EPCAM surface protein expression in cells of an extramedullary lesion of our patient at 6 month follow-up. **B**, Heatmap showing the comparison of expression of selected genes between scRNA-seq data of the extramedullary lesion of our patient at 6 month follow-up and publicly available IMD scRNA-seq datasets. Selected genes are epithelial- (*EPCAM, SFN, KRT8, KRT18*), extracellular matrix interaction- (*ADGRE2*), cell-cell adehsion- (*ITGB7*), as well as angiogenesis-associated (*VEGFA, PTN, ANG*) genes and *TGFA*. Differential expression analysis results between EMD and IMD can be found in **Supplementary Table 2**. Abbreviations: EMD - extramedullary MM disease; IMD - intramedullary MM disease.

## SUPPLEMENTARY TABLES LEGENDS

**Supplementary Table 1: Data quality; Public datasets; QC filtering; Gene signatures. Page 1**, scRNA-seq data quality metrics from CellRanger output of data generated in the present study. ‘Identifier’ shows the name of the library in the manuscript. ‘Library’ shows the formal library name (also used in EGA). **Page 2**, Description of publicly available datasets used for data integration and projection. ‘Reference’ shows an identifier based on the first author of the respective study. ‘Title’ shows the manuscript title. ‘Journal’ indicates where the article was published. ‘Condition’ indicates the underlying disease context. ‘PMID’ and ‘DOI’ show identifiers to access the manuscripts. The database and respective accession numbers (database: accession no.) for the datasets can be found at ‘Access’. **Page 3**, Quality metrics and customized parameters used for quality filtering of all datasets that were used in this study. Parameters were customized in order to account for varying underlying data qualities between studies. ‘Identifier’ indicates the name of the datasets generated in the present study or the first author of the respective study public data is derived from (analogue to ‘Reference’ in Table page 2). ‘Condition’ indicates the underlying disease context. **Page 4**, Description of Th1-polarized Th17 signatures used for module score calculation. ‘Reference’ shows an identifier based on the first author of the respective study. ‘Title’ shows the manuscript title. ‘Cluster’ indicates the subset from the original publication. ‘PMID’ and ‘DOI’ show identifiers to access the manuscripts. ‘Genes’ shows the gene set of the respective signature.

**Supplementary Table 2: Differential expression analysis results. Page 1-11**, Statistics related to results of conducted differential expression analyses. The tables are ordered by cell type (column 1 ‘cluster’). Genes (column 2 for gene symbol & column 8 for ENSEMBL ID) are ordered by increasing p-values (column 6) and decreasing average log-fold change (column 2). Bonferroni-method was used to adjust p-values for multiple comparisons (column 7). Percentages of non-zero expressing cells in the respective ‘cluster’ and all other cells are indicated (column 4 for ‘cluster’ & column 5 for all other cells).

## REFERENCES

1. Rasche, L., Hudecek, M. & Einsele, H. What is the future of immunotherapy in multiple myeloma? Blood 136, 2491–2497 (2020).

2. Munshi, N. C., Hege, K. & San-Miguel, J. Idecabtagene Vicleucel in Relapsed Myeloma. Reply. The New England journal of medicine vol. 384 2357–2358 (2021).

3. Rasche, L. et al. Low expression of hexokinase-2 is associated with false-negative FDG-positron emission tomography in multiple myeloma. Blood 130, 30–34 (2017).

4. Danylesko, I. et al. Immune imitation of tumor progression after anti-CD19 chimeric antigen receptor T cells treatment in aggressive B-cell lymphoma. Bone Marrow Transplant. 56, 1134–1143 (2021).

5. Wang, J. et al. Role of Fluorodeoxyglucose Positron Emission Tomography/Computed Tomography in Predicting the Adverse Effects of Chimeric Antigen Receptor T Cell Therapy in Patients with Non-Hodgkin Lymphoma. Biol. Blood Marrow Transplant. 25, 1092–1098 (2019).

6. Fujimoto, D. et al. Pseudoprogression in Previously Treated Patients with Non-Small Cell Lung Cancer Who Received Nivolumab Monotherapy. J. Thorac. Oncol. 14, 468–474 (2019).

7. Ferrara, R. et al. Hyperprogressive Disease in Patients With Advanced Non–Small Cell Lung Cancer Treated With PD-1/PD-L1 Inhibitors or With Single-Agent Chemotherapy. JAMA Oncology vol. 4 1543 Preprint at https://doi.org/10.1001/jamaoncol.2018.3676 (2018).

8. Gkiozos, I. et al. Sarcoidosis-Like Reactions Induced by Checkpoint Inhibitors. J. Thorac. Oncol. 13, 1076–1082 (2018).

9. Zhou, X. et al. Salvage therapy with ‘Dara-KDT-P(A)CE’ in heavily pretreated, high-risk, proliferative, relapsed/refractory multiple myeloma. Hematol. Oncol. 40, 202–211 (2022).

10. Nobashi, T. et al. The utility of PET/CT with (68)Ga-DOTATOC in sarcoidosis: comparison with (67)Ga-scintigraphy. Ann. Nucl. Med. 30, 544–552 (2016).

11. Lapa, C. et al. [Ga]Pentixafor-PET/CT for imaging of chemokine receptor CXCR4 expression in multiple myeloma - Comparison to [F]FDG and laboratory values. Theranostics 7, 205–212 (2017).

12. Acosta-Rodriguez, E. V. et al. Surface phenotype and antigenic specificity of human interleukin 17–producing T helper memory cells. Nature Immunology vol. 8 639–646 Preprint at https://doi.org/10.1038/ni1467 (2007).

13. Ramesh, R. et al. Pro-inflammatory human Th17 cells selectively express P-glycoprotein and are refractory to glucocorticoids. J. Exp. Med. 211, 89–104 (2014).

14. Kamali, A. N. et al. A role for Th1-like Th17 cells in the pathogenesis of inflammatory and autoimmune disorders. Mol. Immunol. 105, 107–115 (2019).

15. Basdeo, S. A. et al. Ex-Th17 (Nonclassical Th1) Cells Are Functionally Distinct from Classical Th1 and Th17 Cells and Are Not Constrained by Regulatory T Cells. J. Immunol. 198, 2249–2259 (2017).

16. Duhen, T. & Campbell, D. J. IL-1β promotes the differentiation of polyfunctional human CCR6+CXCR3+ Th1/17 cells that are specific for pathogenic and commensal microbes. J. Immunol. 193, 120–129 (2014).

17. Ramstein, J. et al. IFN-γ-Producing T-Helper 17.1 Cells Are Increased in Sarcoidosis and Are More Prevalent than T-Helper Type 1 Cells. Am. J. Respir. Crit. Care Med. 193, 1281–1291 (2016).

18. Cerboni, S., Gehrmann, U., Preite, S. & Mitra, S. Cytokine-regulated Th17 plasticity in human health and diseases. Immunology 163, 3–18 (2021).

19. Broos, C. E. et al. Increased T-helper 17.1 cells in sarcoidosis mediastinal lymph nodes. Eur. Respir. J. 51, (2018).

20. Ciric, B., El-behi, M., Cabrera, R., Zhang, G.-X. & Rostami, A. IL-23 Drives Pathogenic IL-17-Producing CD8+ T Cells. The Journal of Immunology 182, 5296–5305 (2009).

21. Liang, Y., Pan, H.-F. & Ye, D.-Q. Tc17 Cells in Immunity and Systemic Autoimmunity. Int. Rev. Immunol. 34, 318–331 (2015).

22. Henriques, A. et al. Frequency and functional activity of Th17, Tc17 and other T-cell subsets in Systemic Lupus Erythematosus. Cell. Immunol. 264, 97–103 (2010).

23. Henriques, A. et al. Distribution and functional plasticity of peripheral blood Th(c)17 and Th(c)1 in rheumatoid arthritis. Rheumatol. Int. 33, 2093–2099 (2013).

24. Basdeo, S. A. et al. Polyfunctional, Pathogenic CD161+ Th17 Lineage Cells Are Resistant to Regulatory T Cell-Mediated Suppression in the Context of Autoimmunity. J. Immunol. 195, 528–540 (2015).

25. Liao, M. et al. Single-cell landscape of bronchoalveolar immune cells in patients with COVID-19. Nat. Med. 26, 842–844 (2020).

26. Wendisch, D. et al. SARS-CoV-2 infection triggers profibrotic macrophage responses and lung fibrosis. Cell 184, 6243–6261.e27 (2021).

27. Liao, S.-Y. et al. Single-cell RNA sequencing identifies macrophage transcriptional heterogeneities in granulomatous diseases. Eur. Respir. J. 57, (2021).

28. Nathan, A. et al. Multimodally profiling memory T cells from a tuberculosis cohort identifies cell state associations with demographics, environment and disease. Nat. Immunol. 22, 781–793 (2021).

29. Gideon, H. et al. Multimodal profiling of lung granulomas in macaques reveals cellular correlates of tuberculosis control. Immunity 55, 827–846.e10 (2022).

30. Lozupone, C., Lladser, M. E., Knights, D., Stombaugh, J. & Knight, R. UniFrac: an effective distance metric for microbial community comparison. ISME J. 5, 169–172 (2011).

31. Damsky, W. et al. Inhibition of type 1 immunity with tofacitinib is associated with marked improvement in longstanding sarcoidosis. Nat. Commun. 13, 1–17 (2022).

32. Morse, C. et al. Proliferating SPP1/MERTK-expressing macrophages in idiopathic pulmonary fibrosis. Eur. Respir. J. 54, (2019).

33. Da Vià, M. C. et al. Homozygous BCMA gene deletion in response to anti-BCMA CAR T cells in a patient with multiple myeloma. Nat. Med. 27, 616–619 (2021).

34. Jong, M. M. E. de et al. The multiple myeloma microenvironment is defined by an inflammatory stromal cell landscape. Nature Immunology vol. 22 769–780 Preprint at https://doi.org/10.1038/s41590-021-00931-3 (2021).

35. Lapa, C. et al. CXCR4-directed endoradiotherapy induces high response rates in extramedullary relapsed Multiple Myeloma. Theranostics 7, 1589–1597 (2017).

36. van Langelaar, J. et al. T helper 17.1 cells associate with multiple sclerosis disease activity: perspectives for early intervention. Brain 141, 1334–1349 (2018).

37. Dhodapkar, K. M. et al. Dendritic cells mediate the induction of polyfunctional human IL17-producing cells (Th17-1 cells) enriched in the bone marrow of patients with myeloma. Blood 112, 2878–2885 (2008).

38. Lomax, A. J. et al. Immunotherapy-induced sarcoidosis in patients with melanoma treated with PD-1 checkpoint inhibitors: Case series and immunophenotypic analysis. Int. J. Rheum. Dis. 20, 1277–1285 (2017).

39. Axelrod, M. L. et al. T cells specific for α-myosin drive immunotherapy-related myocarditis. Nature 611, 818–826 (2022).

40. Lapa, C. et al. The gross picture: intraindividual tumour heterogeneity in a patient with nonsecretory multiple myeloma. European Journal of Nuclear Medicine and Molecular Imaging vol. 44 1097–1098 Preprint at https://doi.org/10.1007/s00259-017-3656-x (2017).

41. Niemeijer, A.-L., Hoekstra, O. S., Smit, E. F. & de Langen, A. J. Imaging Responses to Immunotherapy with Novel PET Tracers. J. Nucl. Med. 61, 641–642 (2020).

42. Samur, M. K. et al. Biallelic loss of BCMA as a resistance mechanism to CAR T cell therapy in a patient with multiple myeloma. Nat. Commun. 12, 868 (2021).

43. Wang, B. et al. Chimeric Antigen Receptor T Cell Therapy in the Relapsed or Refractory Multiple Myeloma with Extramedullary Disease--a Single Institution Observation in China. Blood vol. 136 6–6 Preprint at https://doi.org/10.1182/blood-2020-140243 (2020).

44. Breeman, W. A. P. et al. Radiolabelling DOTA-peptides with 68Ga. Eur. J. Nucl. Med. Mol. Imaging 32, 478–485 (2005).

45. Werner, R. A. et al. Imaging of Chemokine Receptor 4 Expression in Neuroendocrine Tumors - a Triple Tracer Comparative Approach. Theranostics 7, 1489–1498 (2017).

46. Young, M. D. & Behjati, S. SoupX removes ambient RNA contamination from droplet-based single-cell RNA sequencing data. Gigascience 9, (2020).

47. Stuart, T. et al. Comprehensive Integration of Single-Cell Data. Cell 177, 1888–1902.e21 (2019).

48. McInnes, L., Healy, J., Saul, N. & Großberger, L. UMAP: Uniform Manifold Approximation and Projection. Journal of Open Source Software vol. 3 861 Preprint at https://doi.org/10.21105/joss.00861 (2018).

49. Sikkema, L. et al. An integrated cell atlas of the human lung in health and disease. bioRxiv (2022) doi:10.1101/2022.03.10.483747.

50. Haghverdi, L., Lun, A. T. L., Morgan, M. D. & Marioni, J. C. Batch effects in single-cell RNA-sequencing data are corrected by matching mutual nearest neighbors. Nat. Biotechnol. 36, 421–427 (2018).

51. Luecken, M. D. et al. Benchmarking atlas-level data integration in single-cell genomics. Nat. Methods 19, 41–50 (2021).

52. Liu, Q. et al. Quantitative assessment of cell population diversity in single-cell landscapes. PLoS Biol. 16, e2006687 (2018).

53. LeBlanc, V. G. et al. Single-cell landscapes of primary glioblastomas and matched explants and cell lines show variable retention of inter- and intratumor heterogeneity. Cancer Cell 40, 379–392.e9 (2022).

